# The Effect of Length of Stay in Hospital on Patients’ Health Outcomes: A Quasi-Experimental Study

**DOI:** 10.1101/2024.12.02.24318326

**Authors:** Benedikt Langenberger, Christopher Worsham, Pascal Geldsetzer

**Author notes:** Corresponding author. Address: 1265 Welch Road Stanford, CA 94305.

## Abstract

The causal effect of hospital length of stay on crucial patient outcomes such as readmissions or mortality is under-investigated and therefore unknown for the vast majority of the US population. Existing evidence stems from association studies that are unable to draw causal conclusions. This study leverages Medicare’s two-midnight (2MN) and three-day (3D) rules as two natural experiments to establish causal relationships between hospital length of stay (LOS) and patient outcomes. Using a quasi-experimental regression discontinuity design with data from a large US hospital, we find that the 2MN rule increases LOS by 0.10 days and the 3D rule by 0.21 days, confirming the validity of these rules as instruments for causal inference. However, despite these increases in LOS, there are no significant effects on 90-day mortality or 30-day readmission rates. These findings suggest that while the 2MN and 3D rules effectively extend hospital stays, they do not improve patient-related outcomes, indicating an inefficient use of hospital resources.

---

In 2022, the United States (US) national health expenditures exceeded USD 4.4 trillion, accounting for approximately 17.4 % of the gross domestic product. Of this, hospital expenses accounted for roughly 1.3 trillion, or about 30 % [1]. By 2028, health expenditures are expected to account for over 19.7 % of GDP, with hospital care among the sectors with the highest growth rates, raising the urgent need to control hospital costs [2].

One way to reduce costs would be to prevent unnecessarily long hospital length of stay (LOS). Although hospitalization is often medically necessary, hospitalizations that are longer than they need to be place patients at risk for hospital-acquired complications such as infections, blood clots, falls, depression, reduced mobility, or even death [3, 4, 5]. With each additional day in the hospital carrying both the risk of complications and leading to increased expenses, determining a LOS that maximizes clinical benefit to the patient in the fewest possible days remains a challenge for both clinicians and payors such as Medicare [3].

Unfortunately, examining simple associations between LOS and patient outcomes does little to inform understanding of the *causal* relationship between the two, as there are many confounding factors that lead to both increased LOS and worse clinical outcomes (e.g., age or comorbidities [6, 7]). While adjustment for measurable factors can be done using standard multivariate regression analysis, unmeasured factors will nevertheless bias results in ways that do not permit causal interpretation [8, 9, 10].

However, “natural experiments” created by policies that impact patients in a clinically arbitrary manner offer an opportunity to study *causal* relationships without the necessity of a randomized controlled trial. One such regulation is Medicare’s *two-midnight rule* (2MN rule) that requires beneficiaries to be hospitalized for at least two midnights for hospitals to be paid at the higher level of an inpatient stay (instead of a lower paying observation stay) [11]. From a practical standpoint, this rule impacts the actual LOS of patients depending on whether they were admitted to the hospital before or after midnight: for two otherwise-similar patients to meet criteria for an inpatient stay, the one who was admitted *just after* midnight would have to remain in the hospital almost an entire day longer than the one who was admitted *just before* midnight. Similar considerations exist for Medicare’s *three-day rule* (3D rule) that requires patients be hospitalized for at least three midnights to qualify for post-acute care at a skilled nursing facility (SNF).

Leveraging data from a large tertiary US hospital, we use a quasi-experimental regression discontinuity design (RDD) model to examine whether extending hospital LOS impacts mortality and readmissions by exploiting differences created arbitrarily by the 2MN and 3D rule. We hypothesized that an extra day spent in the hospital, as estimated by our RDD model, does not affect mortality or readmissions, suggesting an inefficient use of hospital resources.

## Leveraging Hospital Routine Data

The study used the Medical Information Mart for Intensive Care IV (MIMIC-IV) dataset [12], which contains detailed electronic health record (EHR) data on all patient stays of patients aged 18 or older at Beth Israel Deaconess Medical Center (BIDMC) between 2008 and 2019 that were treated in an intensive care unit (ICU) or were admitted to the hospital through the emergency department (ED). BIDMC is a major academic teaching hospital for Harvard Medical School located in Boston, Massachusetts with 743 licensed beds [13]. MIMIC-IV includes data on approximately 300,000 patients and their 425,000 emergency department (ED) visits, 430,000 hospital admissions, and 73,000 ICU stays [14]. Death data in the dataset are derived from the Massachusetts State Registry of Vital Records and Statistics [14].

## Sample Restriction for Medicare Rules

We included all patients age 18 and older in the MIMIC-IV databases who were admitted to BIDMC following an ED visit. We excluded patients that were admitted ± 6 minutes around midnight to avoid (non-visually detectable) bias arising from data manipulation around midnight [15].

For the 2MN rule analysis cohort, we narrowed our sample to patients with LOS between 0.5 days (12 hours) and 2.5 days (60 hours) to focus on patients whose LOS could have been plausibly impacted by the 2MN rule. Similarly, for the analysis of the 3D rule cohort, we restricted our analysis to patients with an LOS between 60 hours and 108 hours (i.e., between 2.5 and 4.5 days).

## Outcomes and Covariates

Our exposure of interest was the hospital LOS based on admission and discharge timestamps. Our primary outcome measures were 90-day mortality and 30-day readmission. 90-day mortality was defined as death within 90 days of hospital admission to account for the possibility that the rules may be associated with in-hospital death. Among patients who survived their initial hospitalization, 30-day readmission was defined as a hospitalization at BIDMC post-discharge with an admission date that was between 1 and 30 days after previously being discharged. Same-day readmissions were excluded since it could not be reliably determined whether it was an inpatient transfer or a true readmission. Covariates included age, gender, race, marital status, Clinical Classification Software [16] predicted 90-day mortality, and Charlson weighted index [17, 18], admission year group (2008–2010; 2011–2013; 2014–2016; 2017–2019).

## Deriving Causal Inference from Routine Data

A simple assessment of the causal relationship between hospital LOS and patient outcomes using observational data will be biased by the numerous clinical factors, observed and unobserved, that influence length of stay in a non-random fashion. For example, a patient with a preexisting diagnosis of heart failure who is admitted after a fall at home is at higher risk for both a longer hospital stay and death or readmission. Multivariate adjustment can account for bias introduced by measurable characteristics, but it cannot account for unmeasured characteristics, such as the degree of a patient’s family support at home following hospital discharge. However, the risk of unmeasured confounding can be overcome by taking advantage of situations where a patient’s hospital length of stay is impacted for reasons that are arbitrary with respect to their clinical situation. In the case of the 2MN or the 3D rule, whether a patient is admitted just before or just after midnight is clinically arbitrary, creating a natural experiment by which some patients’ hospitalization is extended in a random fashion.

Regression discontinuity (RDD) design is an established quasi experimental method to draw causal inference from real-world observational data wherein patients’ exposure vary due to a clinically arbitrary cutoff. In a “fuzzy” RDD analysis, where the exposure can be influenced by factors other than the cutoff (e.g., the 2MN or 3D rule is not the sole determinant of LOS), the cutoff functions as an instrumental variable that randomly impacts the *probability* of an additional day in the hospital.

Such an analysis relies on several key assumptions: that the rule impacts the length of stay for otherwise-similar patients admitted just around mid-night (i.e. the ‘first stage’ effect, which can be empirically tested); that the rule impacts patient outcomes solely through its impact on LOS (the exclusion restriction); that there is no outside factor that affects the application of the rule and patient outcomes (the independence assumption); and finally that the rule serves to either increase or decrease LOS, but not both (the monotonicity assumption) [19, 20].

## Medicare Rules may Trigger Discontinuities in Length of Stay

The 2MN rule [21] was introduced in 2013 by the Centers for Medicare and Medicaid Services (CMS) with the intention of removing ambiguity surrounding which hospital-based services could be reimbursed as “inpatient” versus “outpatient” care. Under the rule, “observation” hospitalizations that span less than two midnights are reimbursed as outpatient care through Medicare Part B. Meanwhile, hospitalizations that span two midnights or more are reimbursed as inpatient stays through Medicare Part A at substantially higher rates [11], creating a financial incentive for hospitals to provide care over a longer time period that is clinically disconnected from the Medicare beneficiary’s care needs.

The 3D rule [22] operates similarly, requiring that patients be hospitalized for at least three consecutive days, not including the day of discharge, in order to qualify for discharge to a SNF for rehabilitative care. In effect, a patient’s inpatient hospitalization must therefore span four midnights for the patient to qualify for Medicare to cover SNF services [23], which are often necessary for patients who cannot safely return directly home following an acute illness. Under this rule, Medicare beneficiaries who may be clinically ready for discharge to SNF after, for example, two days of hospitalization must remain in the hospital longer because of the 3D rule, extending their LOS for a clinically-arbitrary reason.

The practical implications of 2MN and 3D rule are most apparent for Medicare beneficiaries who are admitted to the hospital around midnight. For example, consider two similar patients who present to an emergency department with a hip fracture on a Monday night. One happens to be admitted *just before* midnight, while the other is admitted a few minutes later, *just after* midnight, on Tuesday. The first patient will qualify for inpatient reimbursement on Wednesday and for SNF services on Thursday. Meanwhile, the second patient qualifies for inpatient care on Thursday and SNF services on Friday–an additional day of hospitalization despite arriving at, essentially, the same time as the first patient.

Although not all patients are covered by Medicare, we hypothesized that hospital policies and practices may lead to spill-over effects for patients covered by the broad variety of payors [24] [25].

## Comparison of Patients Admitted Before and After Midnight

Characteristics of patients admitted just before compared to just after mid-night were compared using standardized mean-differences (SMD) [26], with an SMD below 0.1 indicating balance with respect to the tested variable [27], as well as for the 2MN and 3D cohorts. We also tested for differences in patient characteristics by estimating the effect of being admitted to the hospital after midnight on patient characteristics using our described RDD estimation strategy [23]. Statistically significant RDD estimates would indicate a discontinuity of the respective covariate around midnight and may be a sign of manipulation.

## Association and Causal Analysis

We began by estimating the non-causal associations between LOS and outcomes using multivariable regression, adjusting for measured clinical characteristics.

We then applied our RDD estimation strategy to derive estimates of the local average treatment effect (LATE), which is the effect of an additional day of hospitalization for patients who would have been discharged sooner *were it not for the Medicare rule*; these patients are known as “compliers” (LATE may also be referred to as the complier average causal effect, or CACE [28]). We also estimated the effect of being admitted after midnight on health outcomes. This effect, which is often referred to as the “reduced form” [29], can be interpreted as the “intention-to-treat” effect [30], i.e., the effect of the policy on health outcomes. In our main analysis, we run the estimations using local polynomial regression, a standard approach for fuzzy RDD [31]. We derived the bandwidth left and right of the midnight cut-off using the optimal bandwidth selection method as described in Calonico et al [32] (see Appendix C for details). We also conducted complier analysis to identify which patients are actually affected by the policies (see Appendix C for details).

## Analyzing Heterogeneity in Treatment Effects

In addition to the overall population in the respective samples affected by the 2MN and the 3D rule, we tested the effect of LOS on health outcomes in several subgroups in order to identify treatment effect heterogeneity.

First, we focused on the subgroups that should be most affected by the two-midnight and the 3D rule, namely Medicare beneficiaries and, for the 2MN rule which was implemented in 2013, for Medicare patients before and after 2013. Next, we analyzed whether heterogeneity exists regarding subgroups that may be subject to discrimination. Here, we focused on heterogeneity by gender (self-reported male vs. female) and race (self-reported white vs. non-white) - two variables known to affect outcomes in healthcare settings [33, 34, 35, 36]. Finally, we report subgroup analysis by chronic comorbidity severity (Charlson weighted index of 0 vs. ≥ 1 [17] and acute illness severity (defined by the Clinical Classification software, CCSR-2023-1 [16]. Here, we assumed that more vulnerable patient groups may less severely be affected by the respective rules because higher medical necessity allows for less arbitrary variation in discharge timing.

Analyses were conducted using R version 4.2.2 and the rdrobust package for RDD [37]. Alpha levels were set at 0.05 (0.025 for each tail). The Institutional Review Boards (IRBs) at both Massachusetts Institute of Technology and the Beth Israel Deaconess Medical Center approved the use of the data for research. The MIMIC-IV (https://mimic-iv.mit.edu) database can be accessed by certified researchers, so no additional informed consent of the patient and ethic approval are required [14].

## Study Population Characteristics

We identified a total of 298,502 hospital admissions that met inclusion criteria from 2008–2019. A histogram of the time of these hospital admissions to an inpatient ward or intensive care unit is displayed in Fig. A1, and the distribution of hospital LOS is displayed in Fig. A2.

Characteristics of patients admitted just before compared to just after midnight for all admitted patients and those in the 2MN and 3D rule cohorts are displayed in Table 1. Apart from marital status, year group and ICU stay for the 3D rule, all SMDs were below 0.1, indicating well-balanced groups and supporting the assumption that midnight divides admitted patients in a clinically arbitrary fashion. Using the RDD approach to measure covariate values around the midnight threshold, we found no statistically significant discontinuity around midnight for either the 2MN or the 3D rule groups, further supporting the assumption of balanced groups on each side of the midnight discontinuity (Table A2; A3).

**Table 1:** Standardized mean differences ± 30 minutes around midnight.

|  | Overall |  |  | 2MN rule |  |  | 3D rule |  |  |
| --- | --- | --- | --- | --- | --- | --- | --- | --- | --- |
|  | Before midnight | After midnight | SMD | Before midnight | After midnight | SMD | Before midnight | After midnight | SMD |
| n | 8518 | 6853 |  | 3190 | 2438 |  | 219 | 186 |  |
| **Sociodemographics** |  |  |  |  |  |  |  |  |  |
| Age (mean (SD)) | 58.29 (19.81) | 58.93 (19.46) | 0.033 | 56.65 (20.07) | 56.19 (20.15) | 0.022 | 78.72 (12.29) | 77.68 (11.60) | 0.087 |
| Gender = M (%) | 4151 (48.7) | 3354 (48.9) | 0.004 | 1490 (46.7) | 1126 (46.2) | 0.010 | 84 (38.4) | 78 (41.9) | 0.073 |
| Race: white (%) | 0.65 (0.48) | 0.67 (0.47) | 0.044 | 0.64 (0.48) | 0.66 (0.48) | 0.036 | 0.79 (0.41) | 0.78 (0.42) | 0.025 |
| Main language=English (%) | 7663 (90.0) | 6161 (89.9) | 0.002 | 2877 (90.2) | 2208 (90.6) | 0.013 | 188 (85.8) | 163 (87.6) | 0.053 |
| Marital status (%) |  |  | 0.050 |  |  | 0.055 |  |  | 0.151 |
| Divorced | 658 ( 7.7) | 497 ( 7.3) |  | 243 ( 7.6) | 162 ( 6.6) |  | 16 ( 7.3) | 17 ( 9.1) |  |
| Married | 2946 (34.6) | 2521 (36.8) |  | 1123 (35.2) | 904 (37.1) |  | 73 (33.3) | 51 (27.4) |  |
| NA | 170 ( 2.0) | 149 ( 2.2) |  | 40 ( 1.3) | 34 ( 1.4) |  | 8 ( 3.7) | 8 ( 4.3) |  |
| Single | 3814 (44.8) | 2948 (43.0) |  | 1456 (45.6) | 1106 (45.4) |  | 65 (29.7) | 54 (29.0) |  |
| Widowed | 930 (10.9) | 738 (10.8) |  | 328 (10.3) | 232 ( 9.5) |  | 57 (26.0) | 56 (30.1) |  |
| Insurance (%) |  |  | 0.041 |  |  | 0.066 |  |  | 0.032 |
| Medicaid | 974 (11.4) | 709 (10.3) |  | 368 (11.5) | 262 (10.7) |  | 7 ( 3.2) | 5 ( 2.7) |  |
| Medicare | 3272 (38.4) | 2598 (37.9) |  | 1147 (36.0) | 816 (33.5) |  | 165 (75.3) | 140 (75.3) |  |
| Other | 4272 (50.2) | 3546 (51.7) |  | 1675 (52.5) | 1360 (55.8) |  | 47 (21.5) | 41 (22.0) |  |
| Year group (%) |  |  | 0.015 |  |  | 0.043 |  |  | 0.168 |
| 2008 - 2010 | 3952 (46.4) | 3179 (46.4) |  | 1548 (48.5) | 1179 (48.4) |  | 104 (47.5) | 103 (55.4) |  |
| 2011 - 2013 | 1924 (22.6) | 1515 (22.1) |  | 705 (22.1) | 552 (22.6) |  | 53 (24.2) | 35 (18.8) |  |
| 2014 - 2016 | 1590 (18.7) | 1312 (19.1) |  | 555 (17.4) | 445 (18.3) |  | 36 (16.4) | 28 (15.1) |  |
| 2017 - 2019 | 1052 (12.4) | 847 (12.4) |  | 382 (12.0) | 262 (10.7) |  | 26 (11.9) | 20 (10.8) |  |
| **Clinical variables** |  |  |  |  |  |  |  |  |  |
| Charlson wscore (mean (SD)) | 1.78 (2.24) | 1.83 (2.23) | 0.023 | 1.33 (1.88) | 1.28 (1.82) | 0.025 | 2.54 (2.14) | 2.55 (2.04) | 0.005 |
| Died in hospital (mean (SD)) | 0.02 (0.13) | 0.02 (0.12) | 0.011 | 0.01 (0.10) | 0.01 (0.09) | 0.013 | 0.00 (0.00) | 0.00 (0.00) | <0.001 |
| ICU stay (mean (SD)) | 0.14 (0.34) | 0.15 (0.35) | 0.026 | 0.04 (0.20) | 0.05 (0.21) | 0.030 | 0.11 (0.31) | 0.14 (0.35) | 0.106 |
| **Treatment and Outcomes** |  |  |  |  |  |  |  |  |  |
| LoS (mean (SD)) | 3.74 (5.50) | 4.25 (6.92) | 0.082 | 1.11 (0.53) | 1.20 (0.53) | 0.167 | 3.22 (0.51) | 3.42 (0.47) | 0.409 |
| 90-day mortality (%) | 591 ( 6.9) | 510 ( 7.4) | 0.020 | 122 ( 3.8) | 88 ( 3.6) | 0.011 | 27 (12.3) | 19 (10.2) | 0.067 |
| 30-day readmission (%) | 1711 (20.3) | 1390 (20.5) | 0.006 | 576 (18.2) | 444 (18.3) | 0.003 | 35 (16.0) | 35 (18.8) | 0.075 |

## Non-causal Association between LOS and Health Outcomes

Non-causal multivariable regression, adjusting for measured covariates, estimated no association between LOS and 90-day mortality in either the 2MN rule or 3D rule cohorts. One additional day of LOS was associated with increased odds of 30-day readmission among patients in the 2MN rule group (adjusted odds ratio 1.062, 95 % CI: 1.031–1.093), but there was no association with readmission among patients included in the 3D rule group (Table A1).

## Causal Effect of Length of Stay on Health Outcomes Triggered by the Two Midnight Rule

The discontinuity in LOS for patients subject to the 2MN rule that were admitted near midnight is displayed in Fig. 1. Patients admitted just before midnight had significantly lower LOS than those admitted just after midnight, with an estimated average increase in LOS of 0.10 days (95 % CI: 0.07–0.13), corresponding to a relative increase of 8.7 % (95 % CI: 6.1 %–11.3 %). Among subgroups, there was a significant increase in LOS observed in each group except Medicare beneficiaries admitted prior to 2013 when the 2MN rule was implemented (Fig. A3).

**Figure 1:**
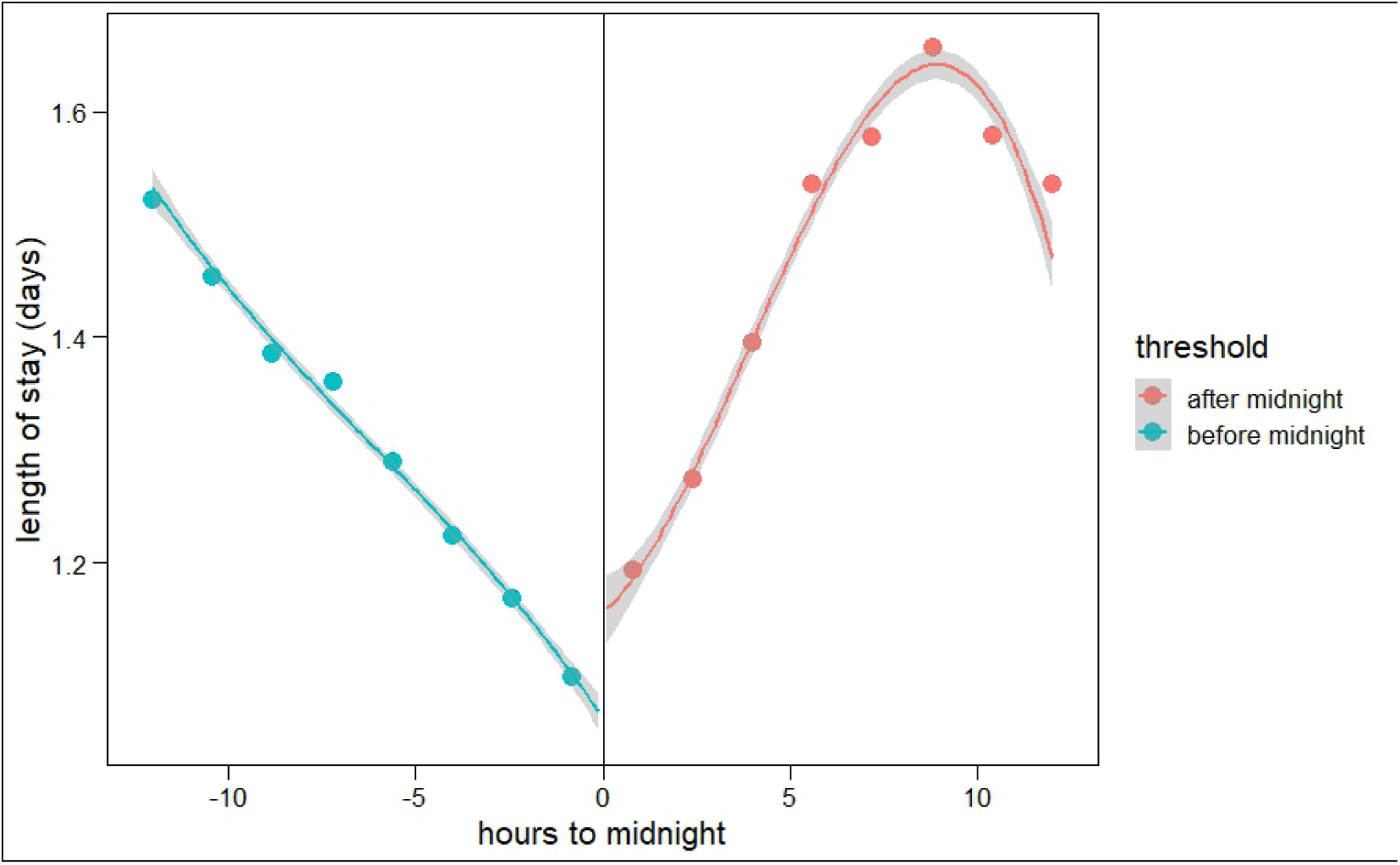
Discontinuity for the two-midnight rule sample *Note*: Discontinuity for the sample that is affected by the 2MN rule (0.5 days to 2.5 days of LOS).

Despite a significant increase in LOS attributable to the 2MN rule, the RDD model estimated no difference in 90-day mortality or 30-day readmission, either overall or among subgroups (Fig. 2; see Fig. for OLS results^1^). As for LOS, we could not detect any statistically significant effect for the ITT analysis for the effect of being admitted after midnight on health outcomes A5.

**Figure 2:**
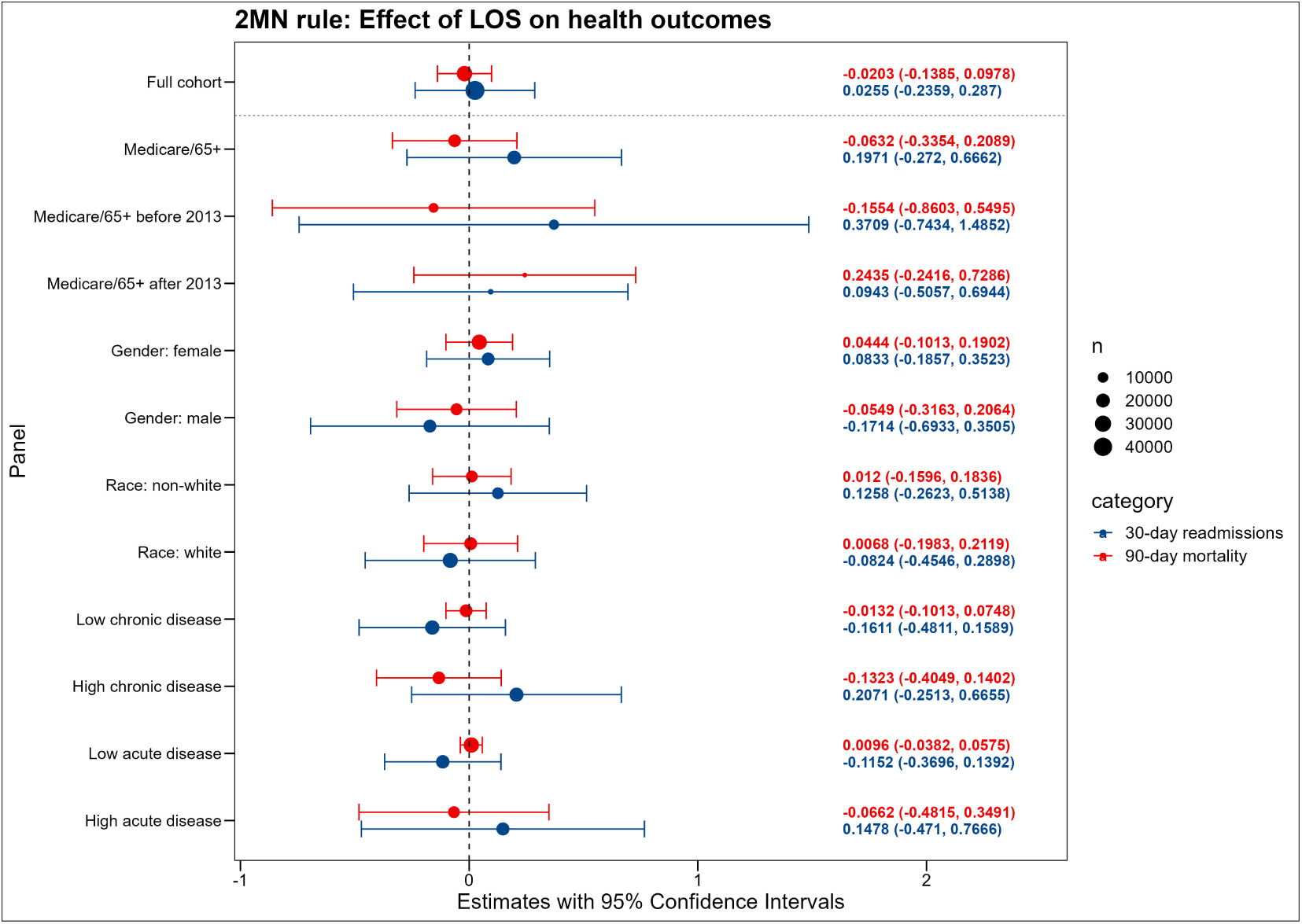
Effect of Length of Stay on Mortality and Readmission for the 2MN rule sample *Note:* Estimated using the local polynomial regression.

## Causal Effect of Length of Stay on Health Outcomes Triggered by the Three Day Rule

The regression discontinuity adjusted LOS for patients subject to the 3D rule that were admitted near midnight are displayed in Fig. 1. Patients admitted just before midnight had significantly lower LOS than those admitted just after midnight, with an estimated average increase in LOS of 0.21 days (95% CI 0.12–0.30), a 6.3% relative increase (95% CI: 3.6 %–9.0 %), attributable to the 3D rule (Fig. A8). Among subgroups, we observed a significant increase in LOS except among non-white patients (Fig. A10).

As before, despite the observed discontinuous increase in LOS, there was no discontinuous difference in 90-day mortality or 30-day readmission either overall or among subgroups. The same holds for the ITT effect, where we could not detect any statistically significant effect of being admitted after midnight A10.

## Differences in LOS by Gender, but not by Race, Were Explained by Mediating Variables

As a result of the heterogeneity by subgroup as seen in Fig. A3, A8, we ran analysis using interaction terms to identify where the heterogeneity for the effect of the two rules on LOS arises for race and gender.

**Figure 3:**
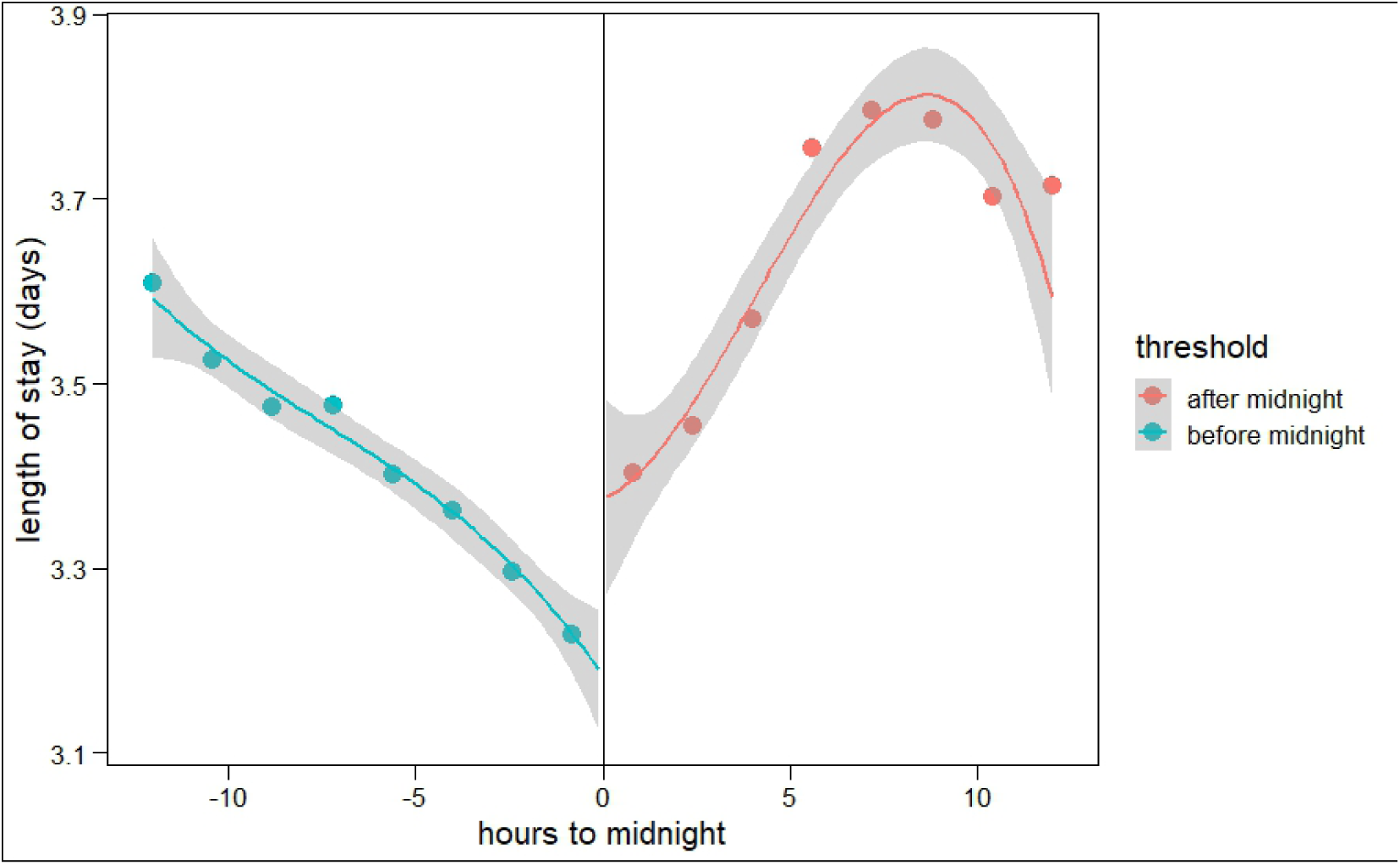
Discontinuity for the three-day rule sample *Note*: Discontinuity for the sample that is affected by the 3D rule. Bandwidth was set to 3 hours.

**Figure 4:**
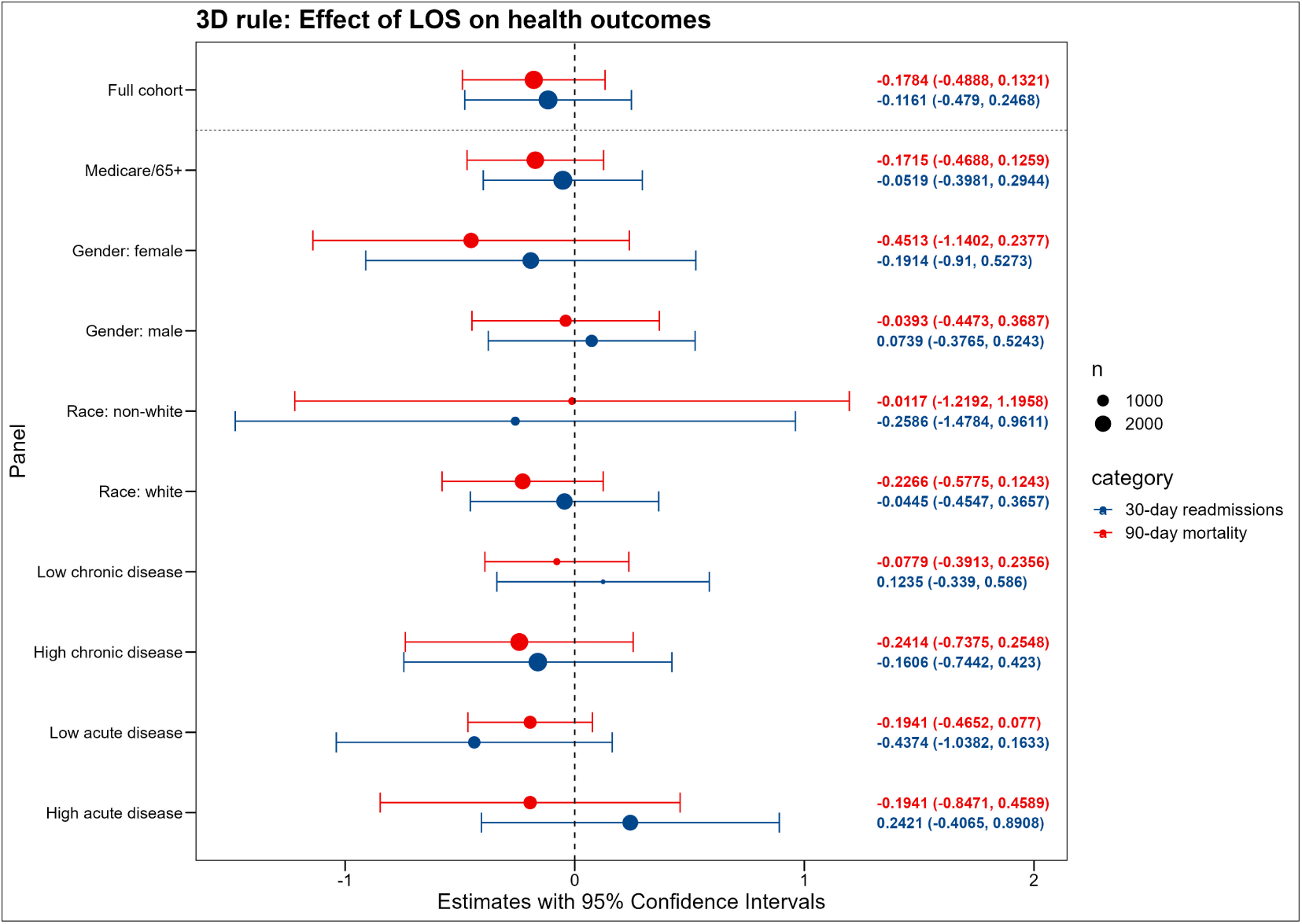
Effect of Length of stay on Mortalit and Readmissions for the three-day rule sample *Note:* Estimated using the local polynomial regression. OLS estimates are presented in the appendix.

We find that these analyses were not able to explain the subgroup heterogeneity by race, but we were able to identify that the heterogeneity by gender was explained by other factors. For the 3D rule, we also found heterogeneity by race and gender, and additional analysis revealed that these effects were mediated by observable variables (see C for details).

## Discussion

In this study of patients admitted to a large, tertiary academic medical center, we observed that despite a clinically arbitrary increase in hospital length of stay attributable to Medicare’s 2-midnight and 3-day rules, there was no difference in 90-day mortality or 30-day readmission rates. These results raise questions about current Medicare regulations that affect healthcare resource utilization without improving outcomes for patients.

Prior observational studies have found both positive and negative non-causal associations of LOS with readmission or mortality [10, 38, 39, 40], leading to speculation about potential causal effects. While one prior study leveraged another midnight rule impacting newborns [15], to our knowledge, this is the first study describing the impact of the 2MN and 3D rules on general adult patient outcomes using quasi-experimental methods that address unmeasured confounding and permit causal inference. Both the newborn study and our study found that additional LOS brought on by arbitrary rules did not impact meaningful patient-centered outcomes. Our findings of an average additional 0.10 days of LOS attributable to the 2MN rule and an additional 0.21 days translate to approximately one in ten patients in the 2MN rule short hospitalization cohort (37.2 % of all hospital patients), as well as one in five patients in the 3D rule longer-hospitalization cohort (2.9 % of all hospital patients) admitted near midnight spending an entire additional day in the hospital for no benefit in mortality or readmission.

Unnecessarily prolonged hospitalizations can lead to myriad costs and problems for patients, hospitals, and payors. Patients do not return home as soon as they could, leaving them away from family and other supports that may be better able to assist in their recovery at home. Financial costs of an additional day of hospitalization are incurred by payors and patients alike. Patients remaining in the hospital unnecessarily also effectively reduce inpatient capacity, which, when strained, can have wide-ranging externalities that negatively impact the care of other hospitalized patients [41]. Although the 2MN and 3D rules are Medicare regulations, the effects of the rules on LOS were similar in magnitude among Medicare beneficiaries as for the entire cohorts which included other payors. This suggests that global hospital practices surrounding the determination of LOS are influenced by Medicare regulations, that other payors have adopted similar regulations, or both.

We found somewhat stronger effects for the 2MN rule for the subgroups female, non-white and low chronic/acute disease patients, compared to their counterparts (Fig.A3). Heterogeneity by race and gender raised the need for further investigation, especially since discrimination based on gender or race is known from other contexts in healthcare [33, 34, 35, 36]. As a result, we identified only unexplained heterogeneity by race for the 2MN rule, which warrants further investigation.

This study comes with some limitations. First, we included patients from all types of insurance and we were limited in distinguishing private insurance/self-payers from Medicare advantage. However, the advantage of this approach was that we were able to determine the policy effects on the overall hospital population in contrast to insurance-type dependent subpopulations. Further, we aimed to disentangle at least the effect for classic Medicare in the subgroup analysis. Second, confidence intervals for the analysis of LOS on health outcomes were broad, reflecting limited sample size around the midnight discontinuity. Third, while we were able to provide real-world evidence with generally good external validity as compared to randomized controlled trials [42], the main limitation here is that we only measure the effects for patients close to an admission time at midnight. We can therefore not necessarily say that these results hold for patients with an admission time further away from midnight as well. Especially for the 3D rule, where we restricted our sample also to patients that were discharged to an SNF, only a small percentage of hospital patients is affected (i.e., the ones that are discharged to an SNF). Fourth, using the RDD approach only allows us to infer the impact of the 2MN and 3D rule around midnight, not necessarily how it affects patients admitted at other times (e.g., admitted at 6pm). It may be plausible to assume that even patients admitted in the late afternoon or early morning are affected by the respective rules, yet it was not possible to test this using our study design. Fifth, we found that the effect of the 2MN rule prior to 2013 was statistically insignificant, although not far from being significant (Fig. A3). Including patients with a weaker effect may have potentially biased our estimates downwards. However, we found a strong overall effect for the full sample, limiting this limitation. Finally, in 2013, a lawsuit against BIDMC was settled due to irregularities in the hospital’s approach to bill Medicare patients regarding inpatient/outpatient stay between 2004 and 2008 [43]. It may be that this lawsuit yields consequences for the effect of the midnight discontinuity before and after 2013. For example, the hospital may have acted more stringently than other hospitals in complying with the 2MN rule following 2013.

## Conclusion

Our study demonstrates that patients affected by the 2MN and the 3D rule experience longer LOS, on average. Per intention-to-treat analysis, we do not find that the policies lead to different health outcomes. Further, the increase in LOS does not have a causal effect on health outcomes. However, heterogeneity regarding the effect of the midnight discontinuity on LOS exists, with non-white patients being disproportionately affected by the rule. Overall, we suggest redesigning the 2MN and the 3D rule as the current state leads to a waste of limited resources that could be more efficiently allocated.

## Data Availability

The Medical Information Mart for Intensive Care IV (MIMIC-IV) is a publicly accessible, deidentified database encompassing detailed health-related data from patients admitted to the intensive care units (ICUs) and emergency departments of the Beth Israel Deaconess Medical Center in Boston, Massachusetts. The database is designed to support a wide array of research studies and educational initiatives, thereby reducing barriers to conducting clinical research.

https://physionet.org/content/mimiciv/2.2/

## Supplementary Materials for

The PDF file includes:

- Figures A1 to A20
- Tables A1 to A8
- Additional text (methods, results and discussion as referenced in the main text)

## A Appendix

### A.1 Figures

**Appendix Figure A1:**
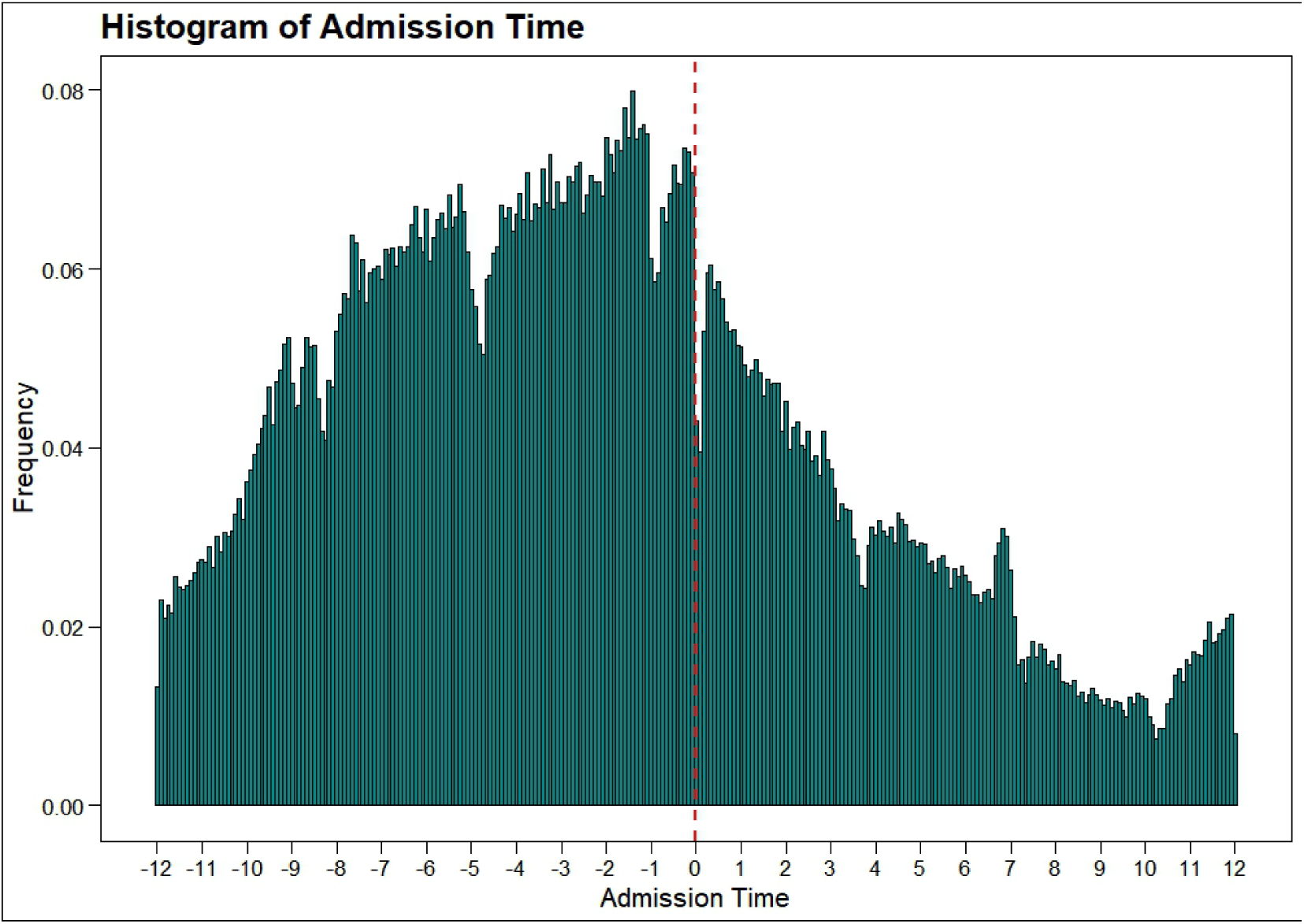
Admission time distribution for patients that came to the hospital through the ED *Note:* Patients admitted ± 6 minutes around midnight were excluded to prohibit any potential bias. Bin width is 5 minutes.

**Appendix Figure A2:**
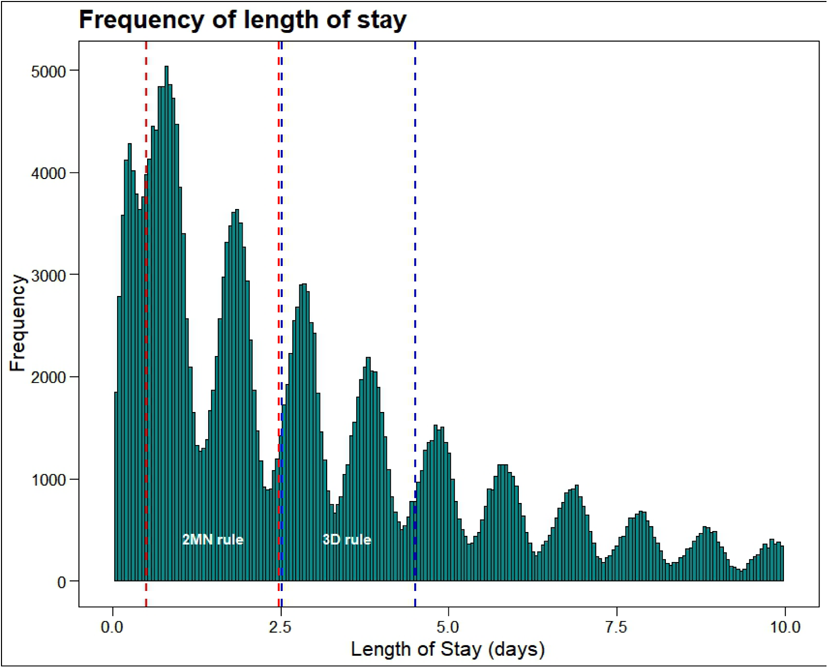
Distribution of total hospital length of stay *Note:* Vertical lines note the boundaries of patients included for analysis of the 2-midnight rule (red) and 3-day rule (blue).

**Appendix Table A1:** Association of LOS with 90-day mortality and 30-day readmission under 2MNR and 3D rule.

|  | 2MN rule |  | 3D rule |  |
| --- | --- | --- | --- | --- |
|  | 90-day mortality | 30-day readmission | 90-day mortality | 30-day readmission |
| Length of Stay | 1.037<br>(0.030) | 1.062***<br>(0.015) | 1.035<br>(0.063) | 0.980<br>(0.053) |
| Constant | -2.839***<br>(0.343) | -0.808***<br>(0.201) | -3.935***<br>(0.742) | 0.730<br>(0.730) |
| Observations | 109,816 | 109,069 | 8,641 | 8,641 |
| Log Likelihood | -14,573.150 | -48,656.440 | -3,032.698 | -3,990.451 |
| Akaike Inf. Crit. | 29,240.300 | 97,406.870 | 6,109.397 | 8,024.902 |

Note: Coefficients are presented as odds ratios. Controlled for age, gender, marital status, Charlson weighted index, anchor year group and race.

**Appendix Figure A3:**
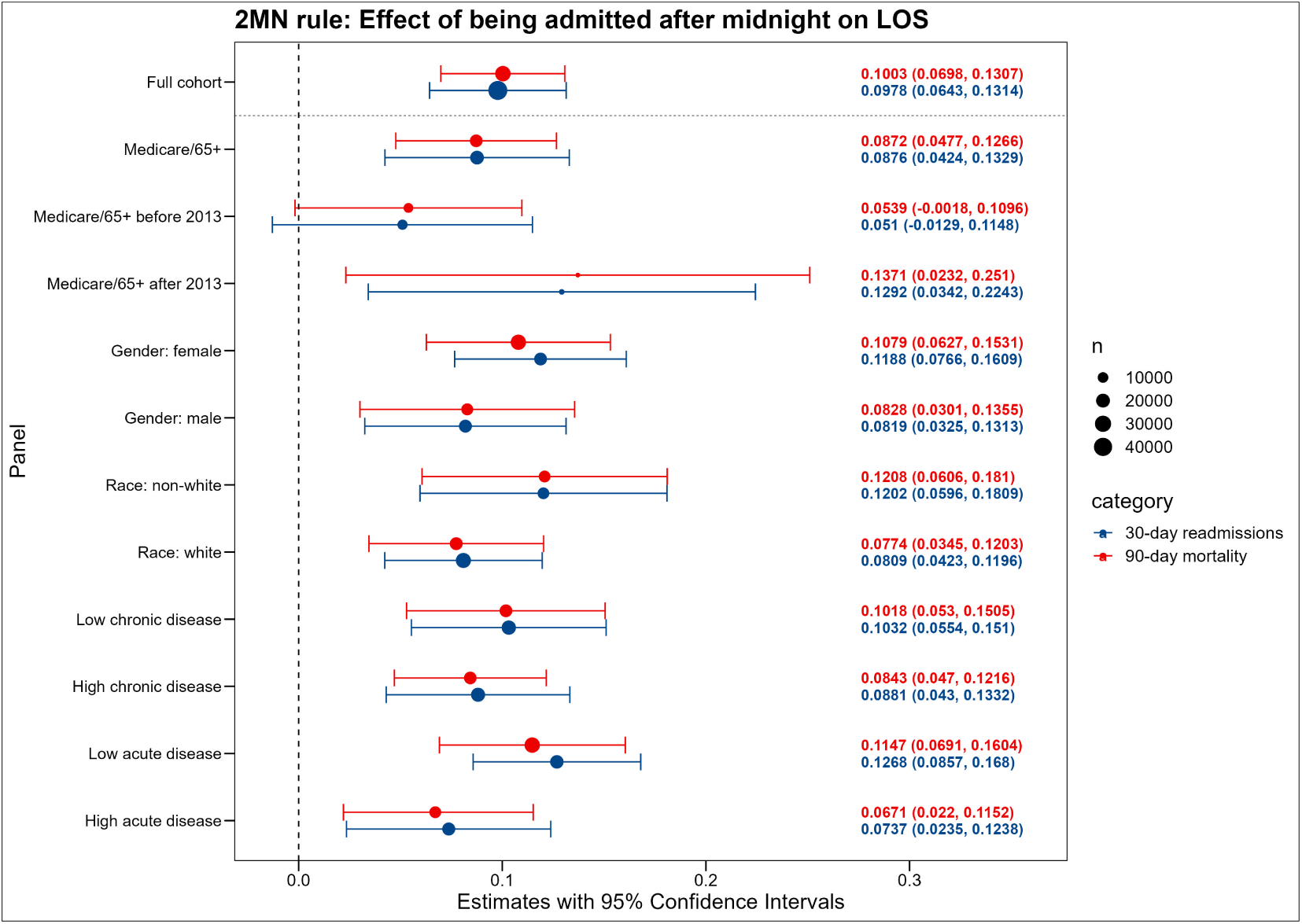
Effect of being admitted after midnight on LOS for the 2MN rule sample *Note:* Estimated using the local polynomial regression.

**Appendix Figure A4:**
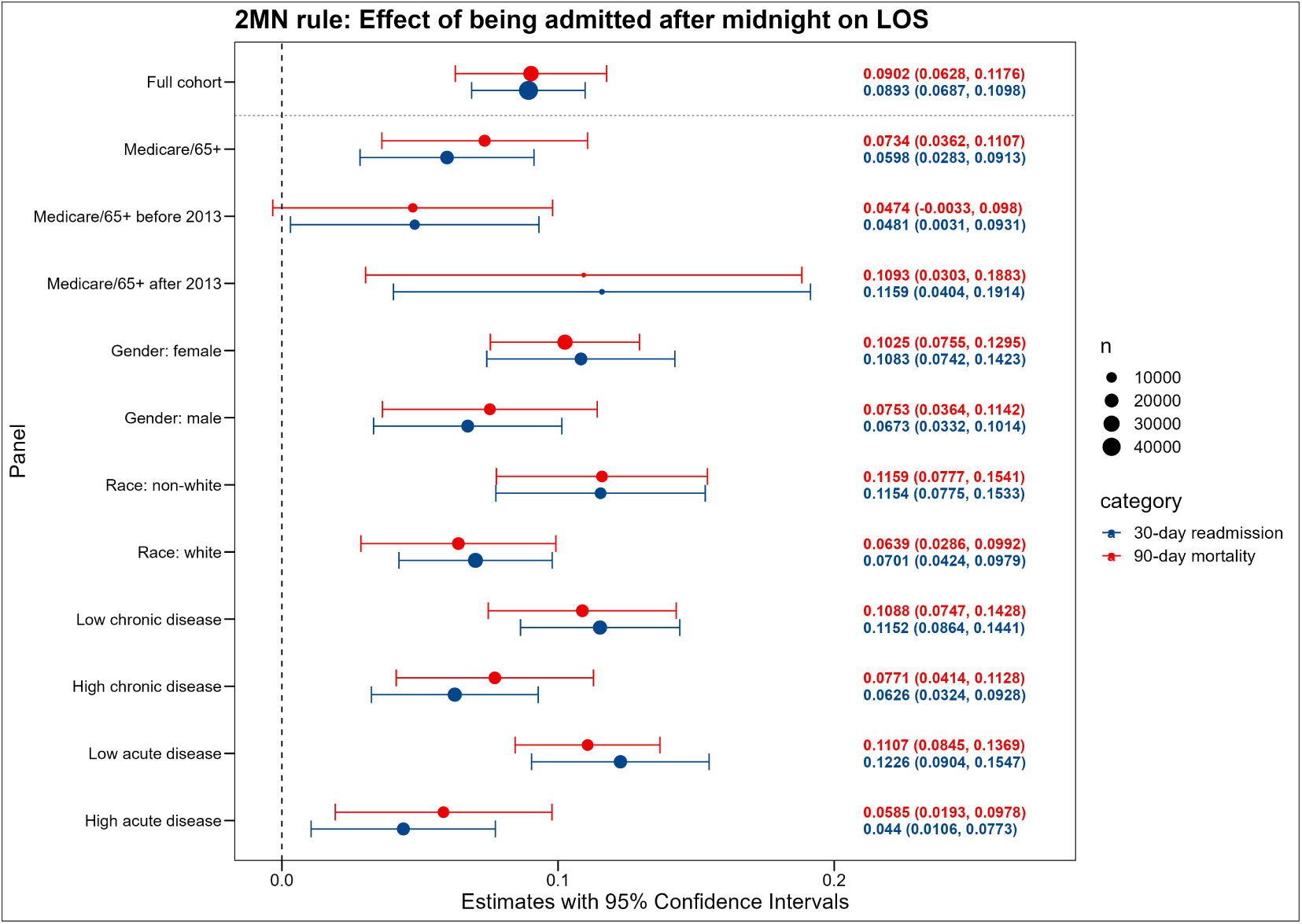
Effect of being admitted after midnight on LOS for the 2MN rule sample *Note:* Estimated using OLS regression.

**Appendix Figure A5:**
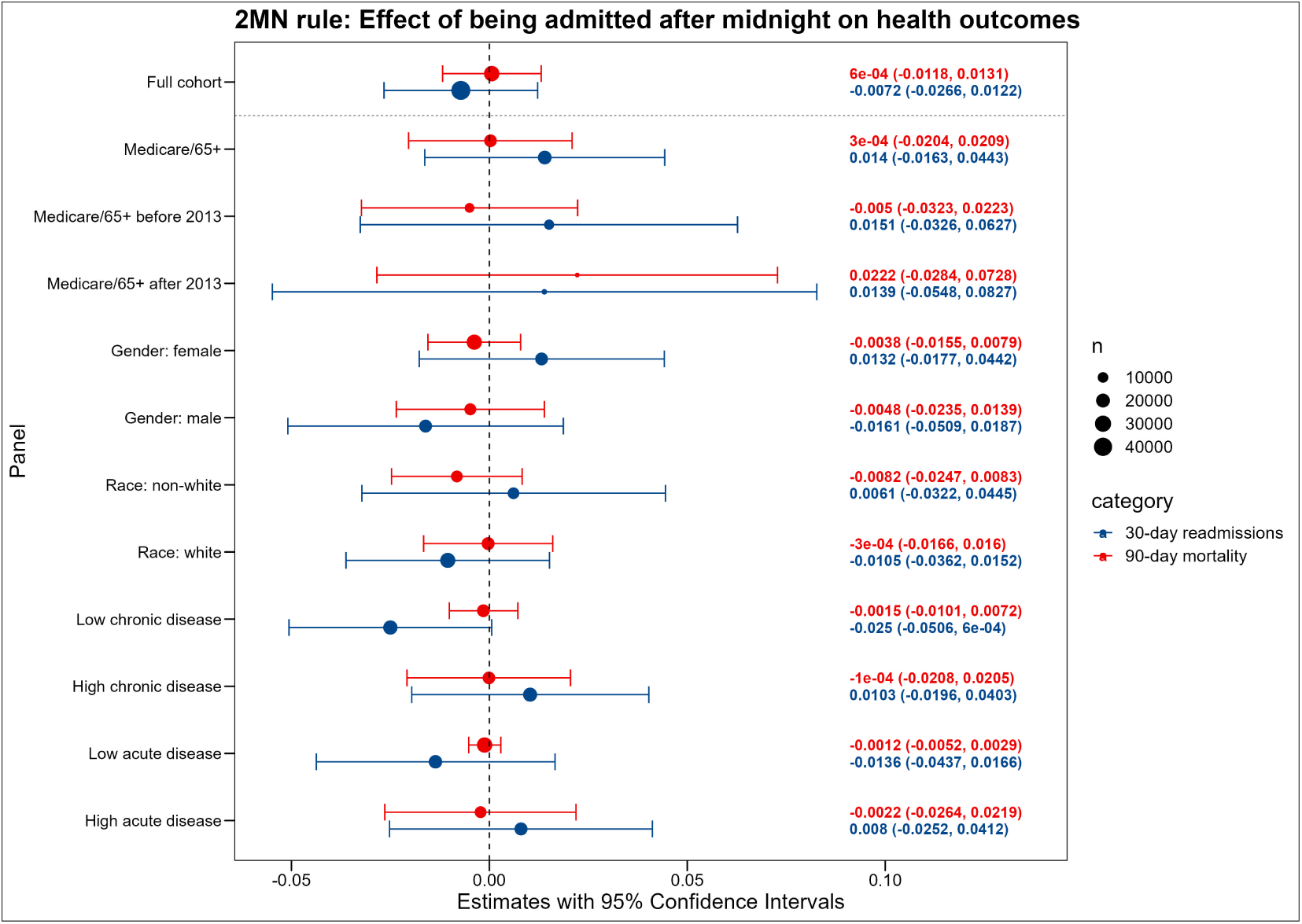
Effect of being admitted after midnight on health outcomes for the 2MN rule sample *Note:* Estimated using local polyomial regression.

**Appendix Figure A6:**
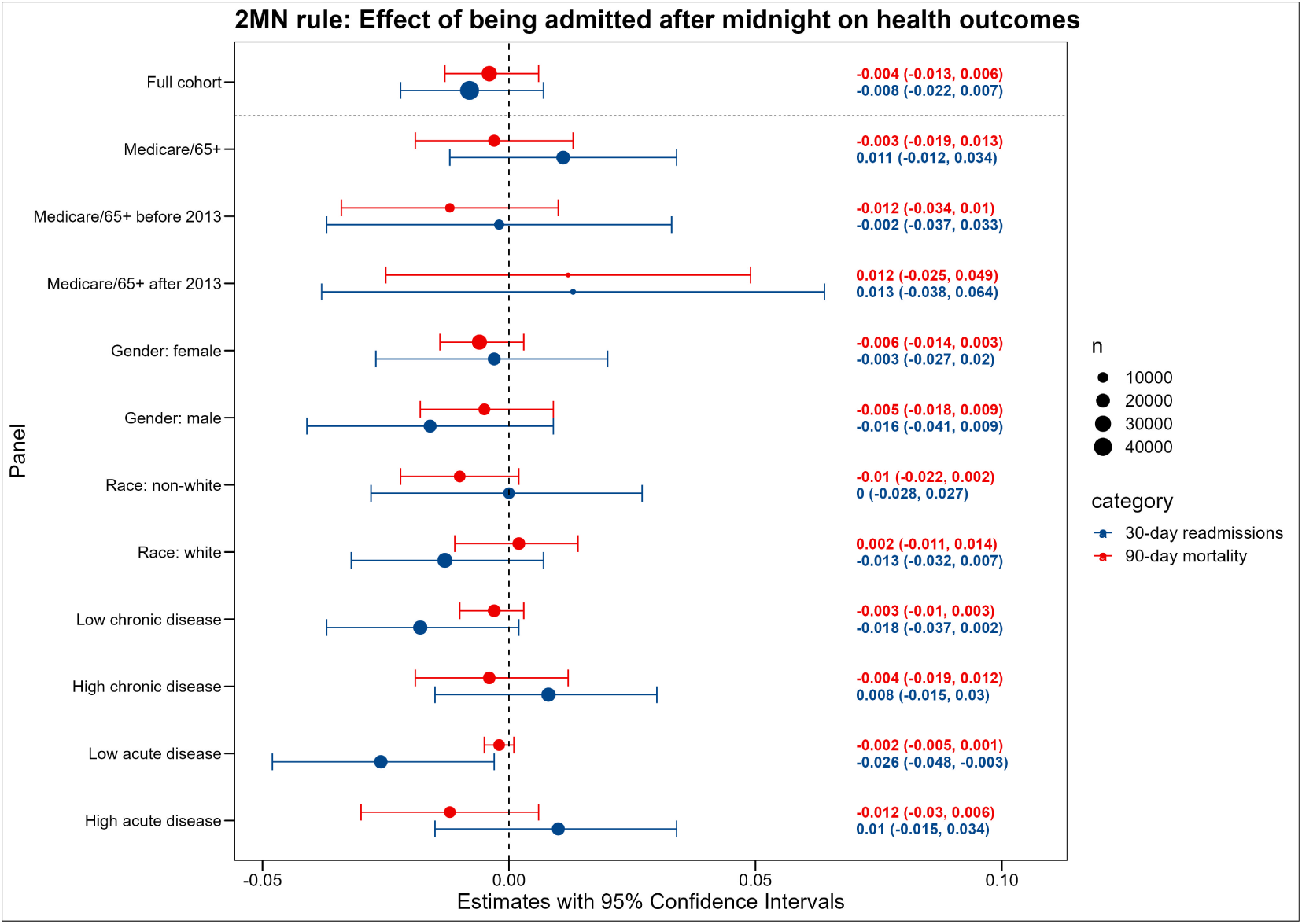
Effect of being admitted after midnight on health outcomes for the 2MN rule sample *Note:* Estimated using OLS regression.

**Appendix Figure A7:**
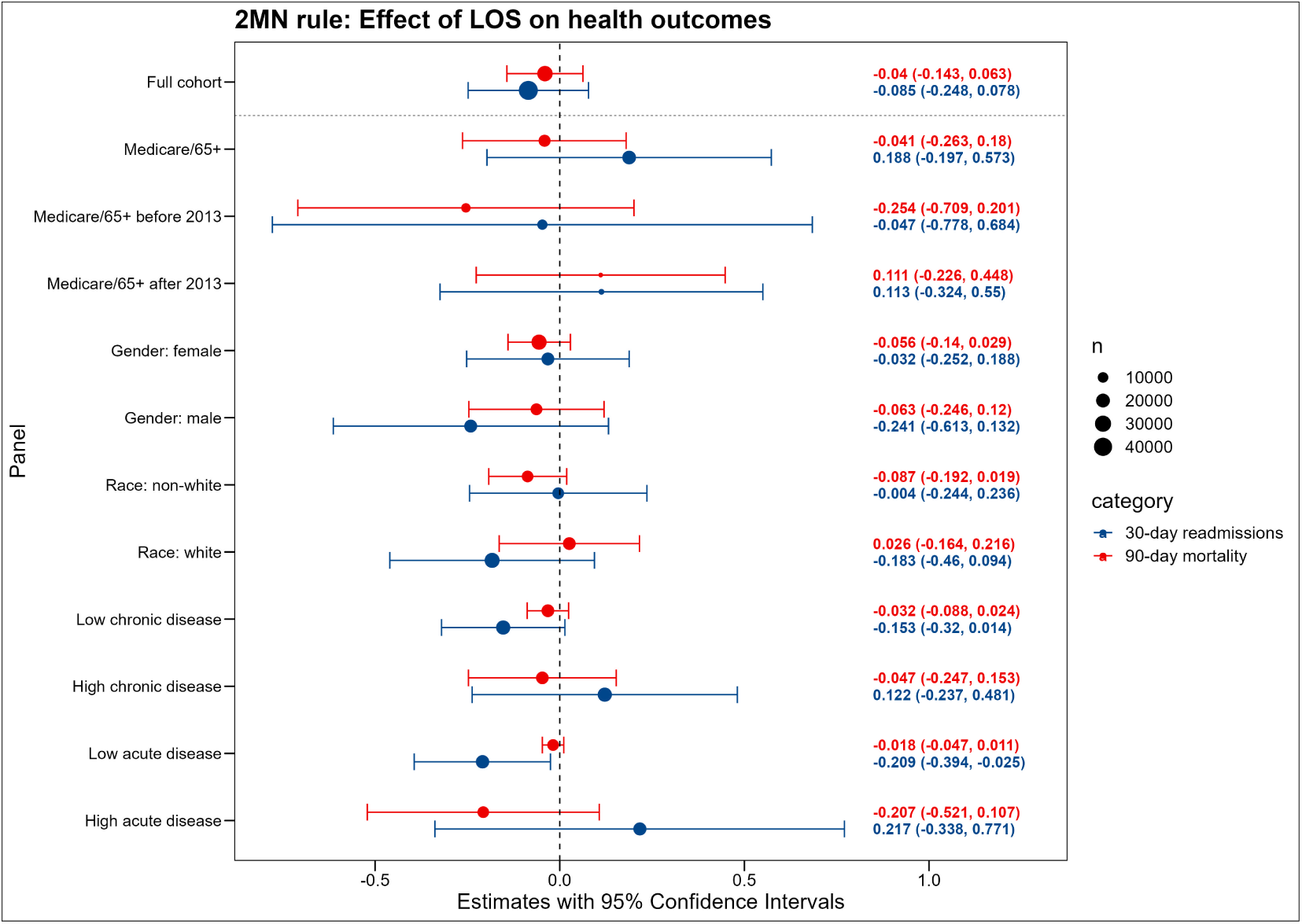
Effect of being admitted after midnight on health outcomes for the 2MN rule sample *Note:* Estimated using OLS regression.

**Appendix Figure A8:**
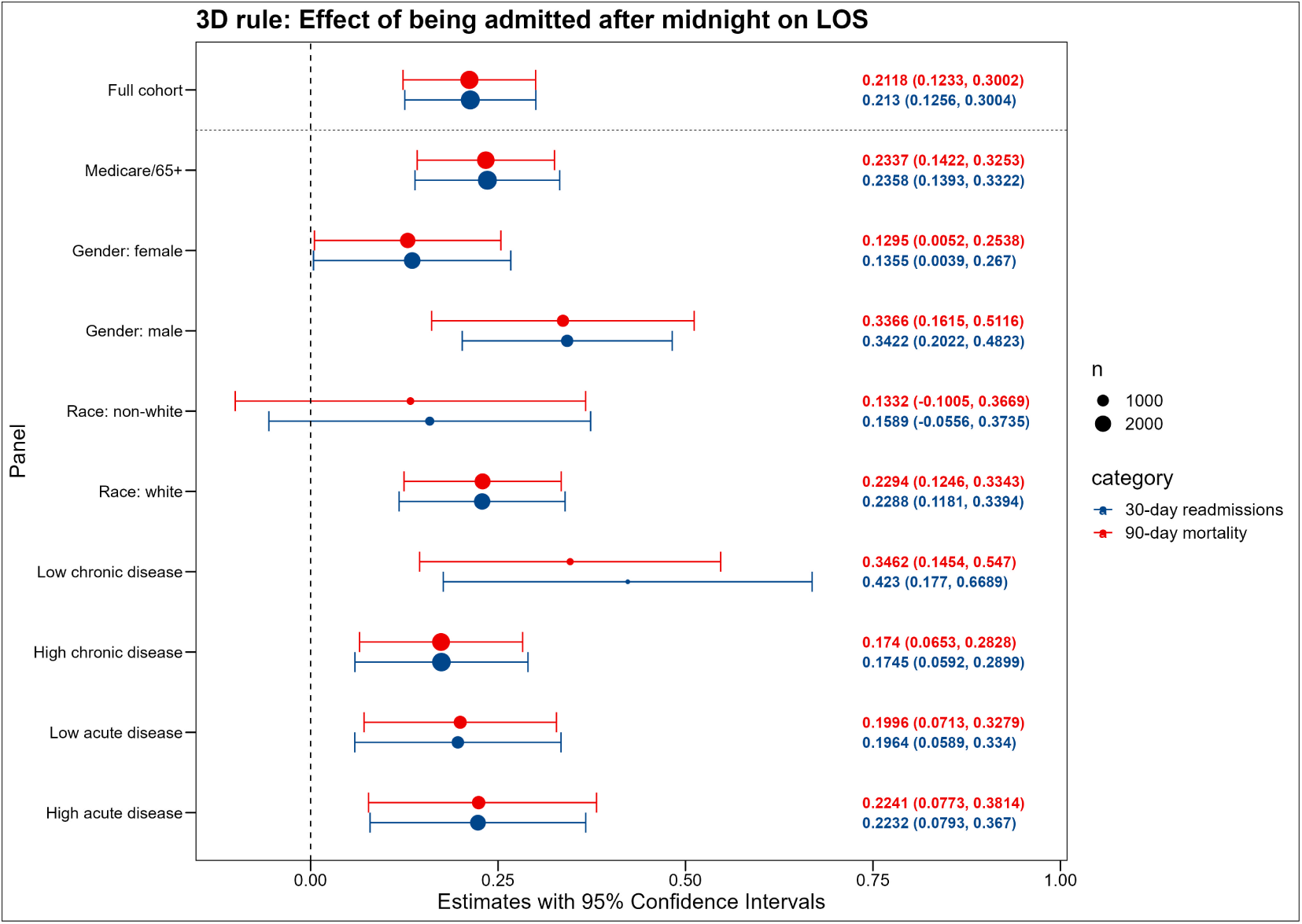
Effect of being admitted after midnight on LOS for the 3D rule sample *Note:* Estimated using local polyomial regression.

**Appendix Figure A9:**
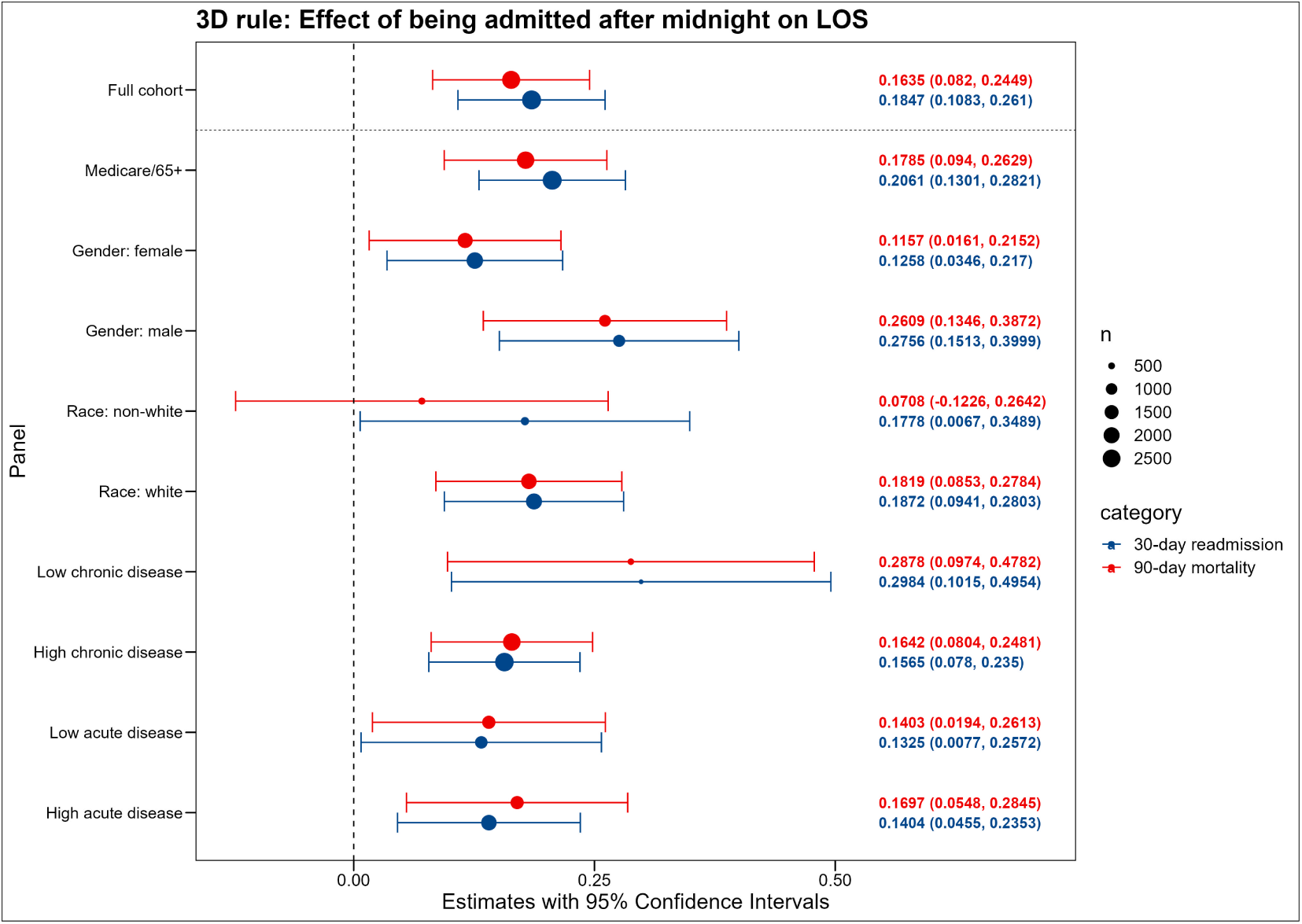
Effect of being admitted after midnight on LOS for the 3D rule sample *Note:* Estimated using OLS regression.

**Appendix Figure A10:**
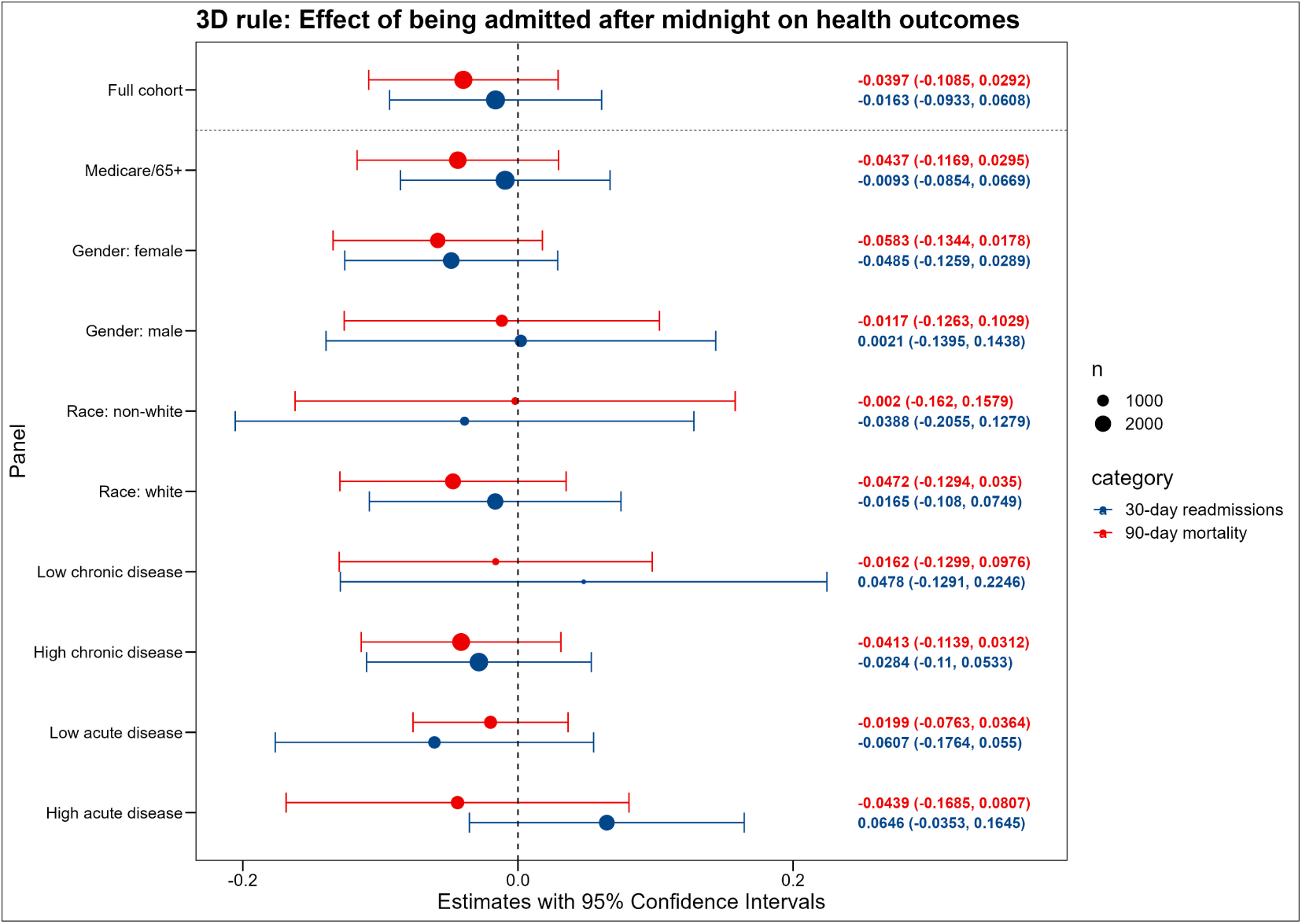
Effect of being admitted after midnight on LOS for the 3D rule sample *Note:* Estimated using local polyomial regression.

**Appendix Figure A11:**
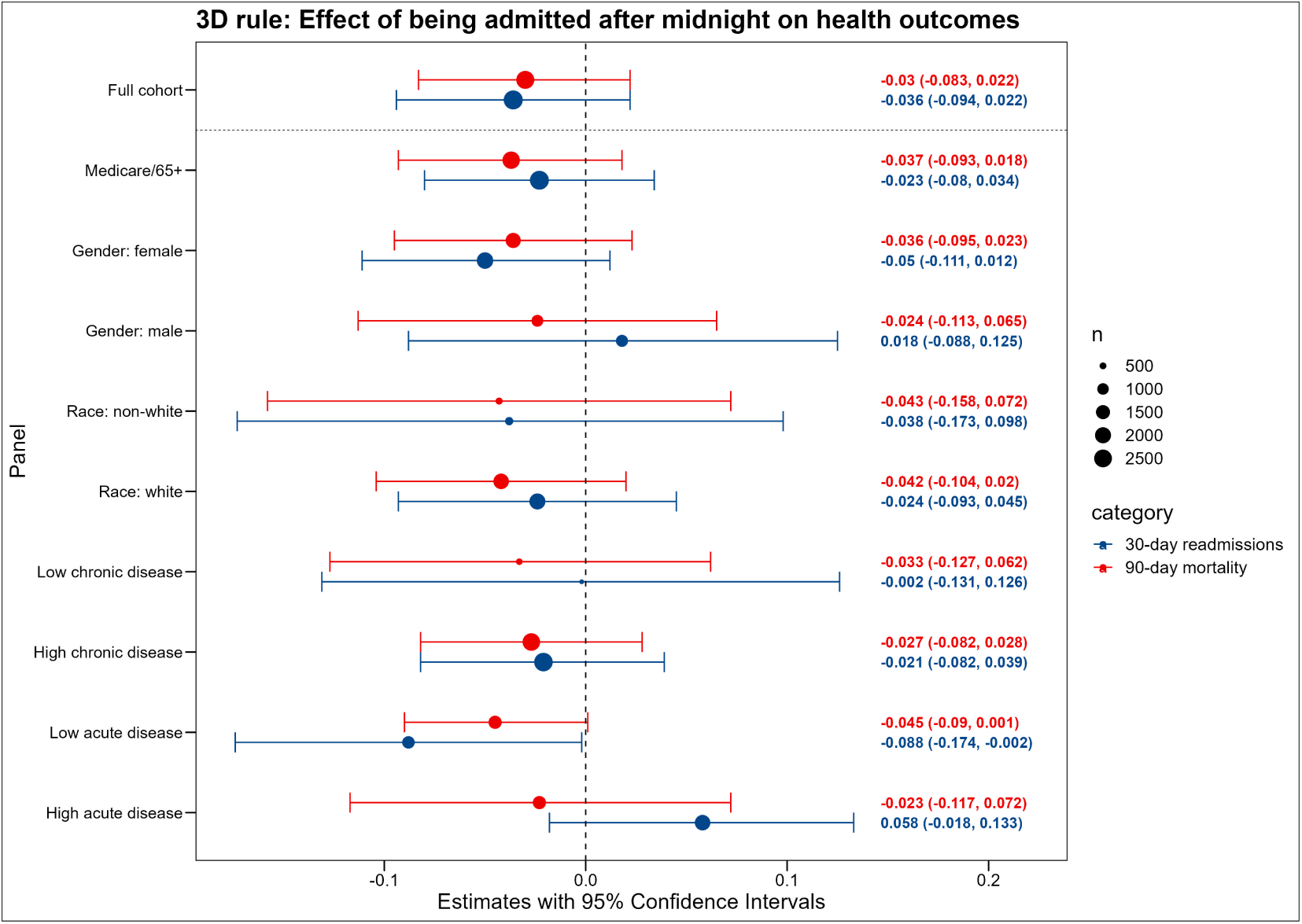
Effect of being admitted after midnight on LOS for the 3D rule sample *Note:* Estimated using OLS regression.

**Appendix Figure A12:**
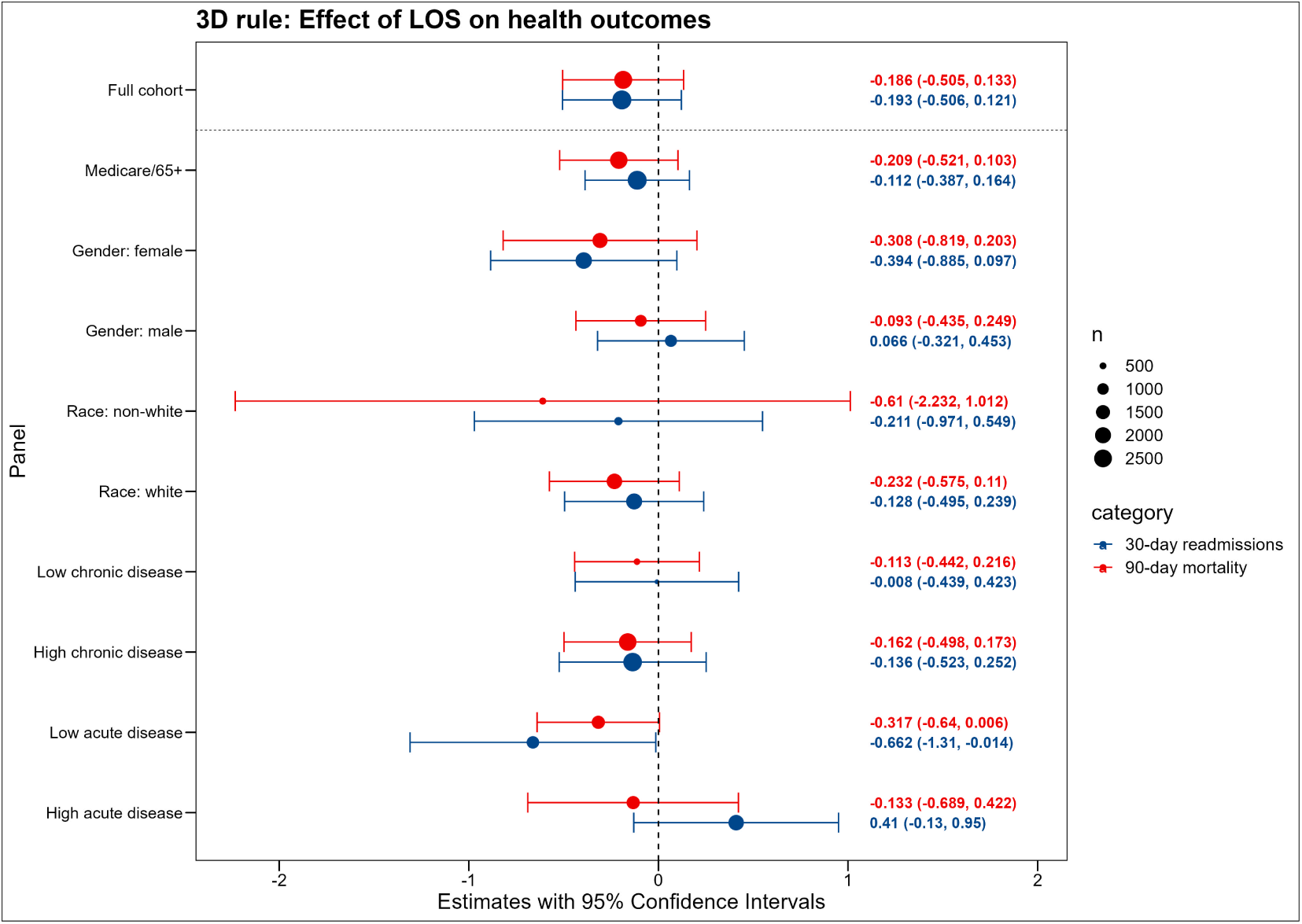
Effect of being admitted after midnight on health outcomes for the 3D rule sample *Note:* Estimated using OLS regression.

**Appendix Figure A13:**
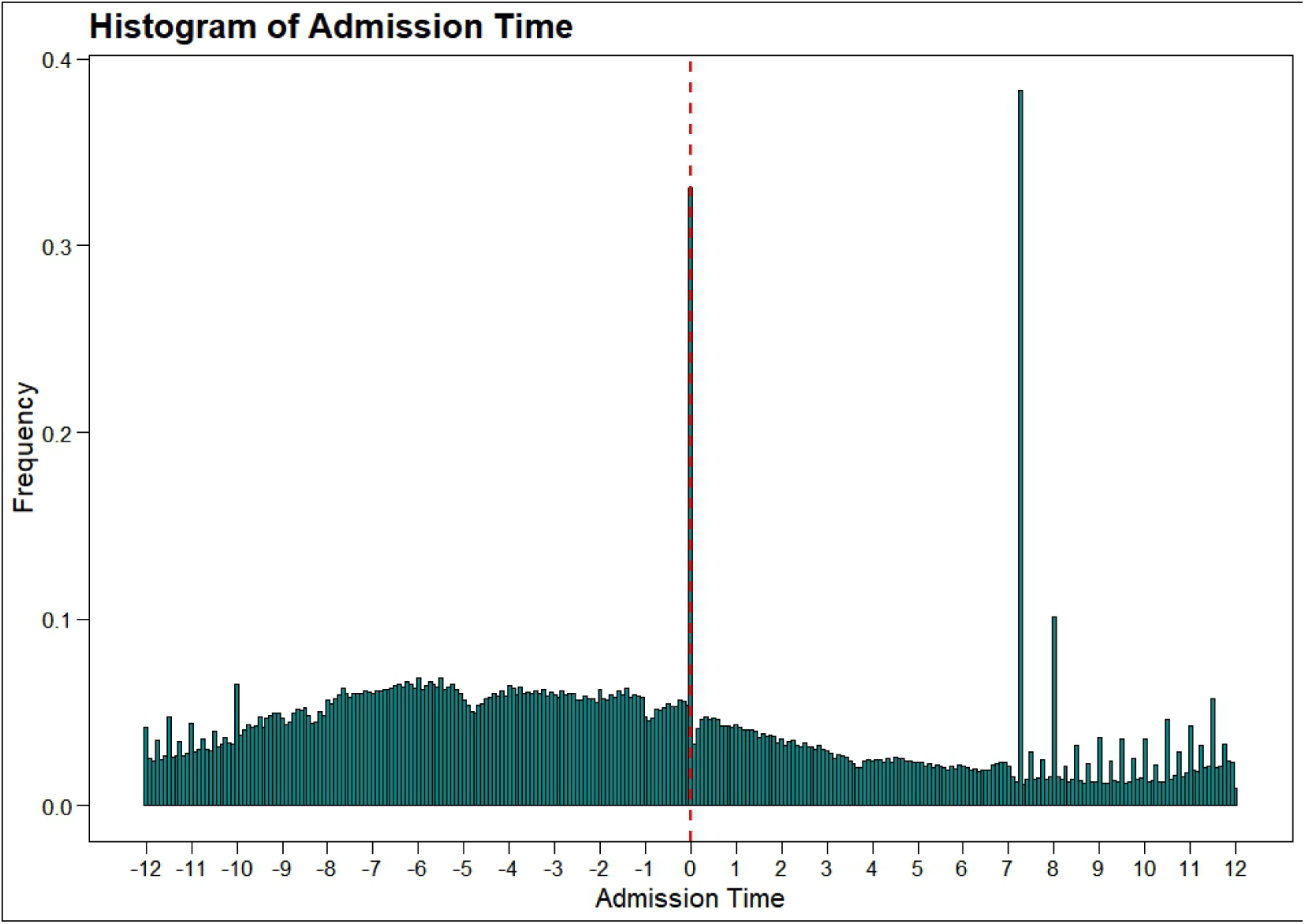
Admission type distribution for all patients including patients that came not through the ED

**Appendix Figure A14:**
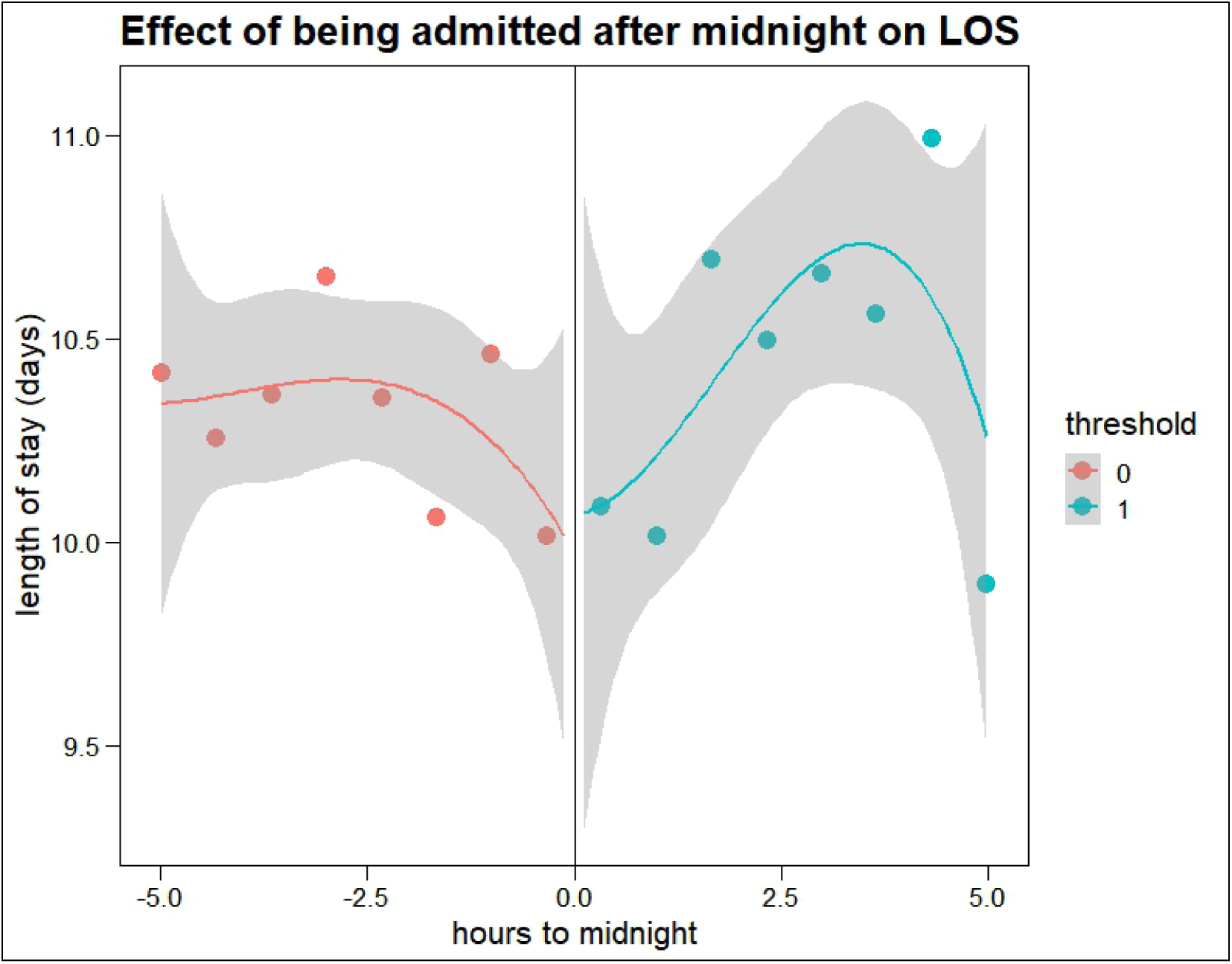
Effect of being admitted after midnight on LOS for patients with LOS *>* 4.5 days. *Note:* Estimated using OLS regression.

**Appendix Figure A15:**
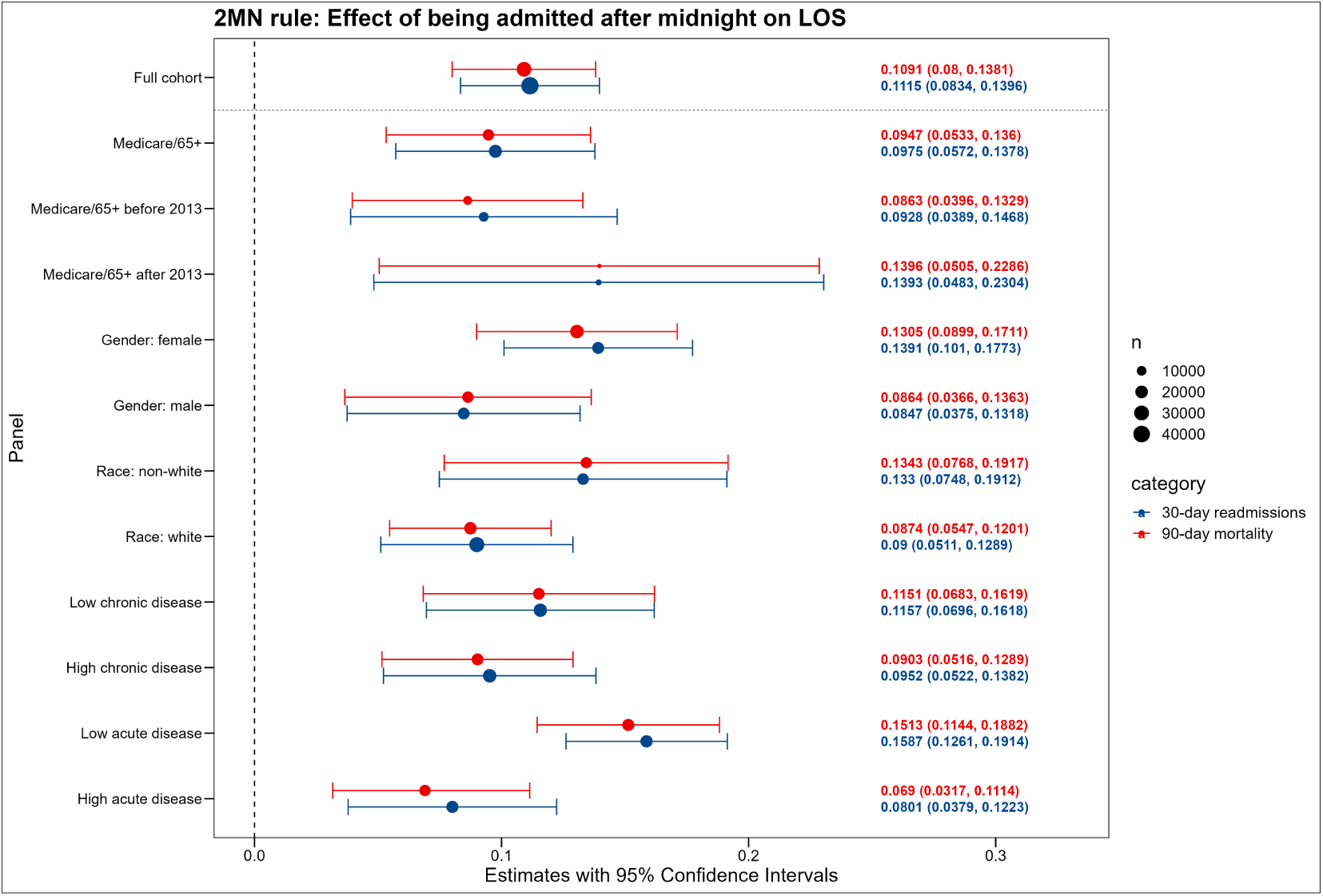
Effect of being admitted after midnight on LOS for the 2MN rule sample *Note:* Estimated using local polyomial regression. Sample without data bunching exclusion.

**Appendix Figure A16:**
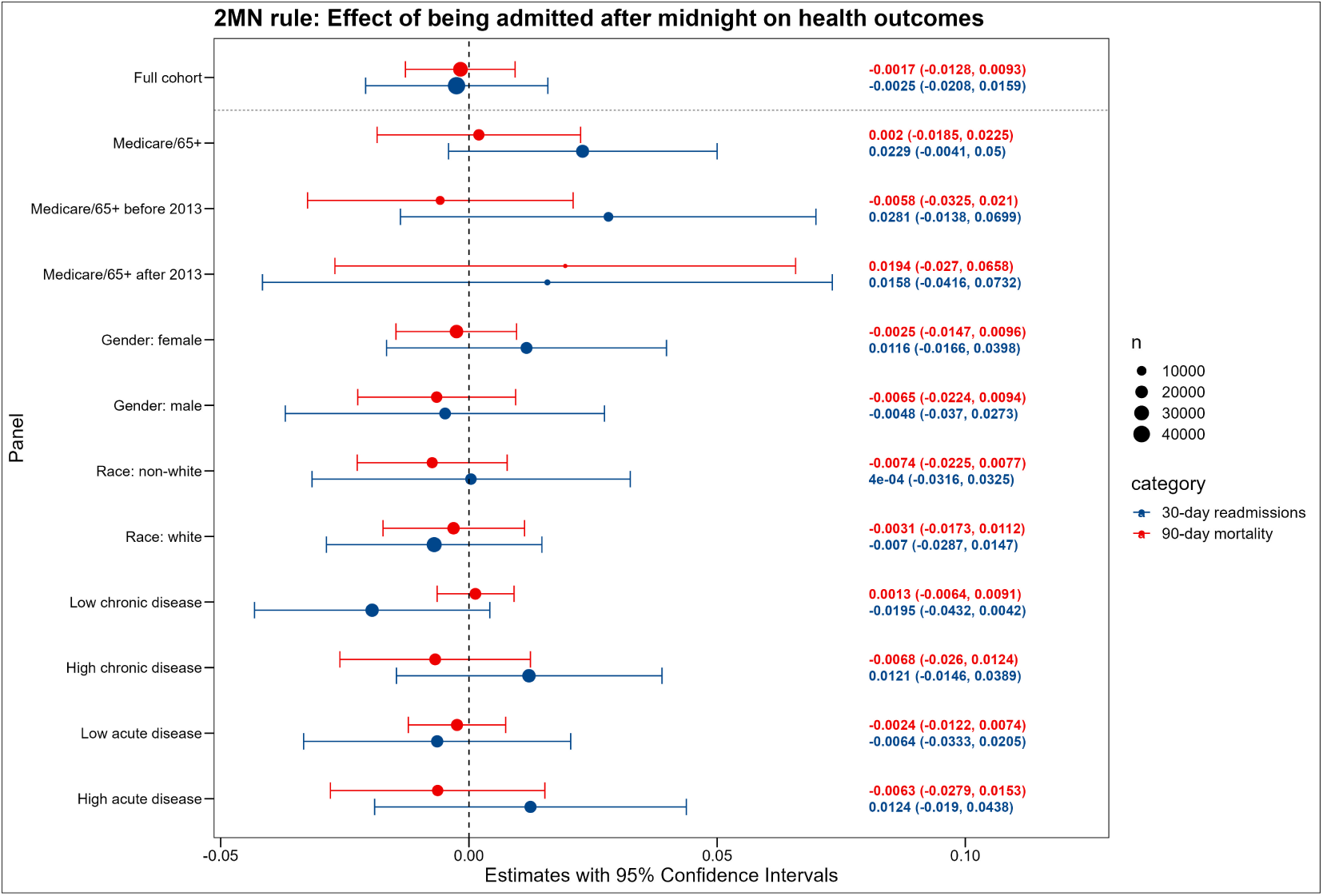
Effect of being admitted after midnight on health outcomes for the 2MN rule sample *Note:* Estimated using local polyomial regression. Sample without data bunching exclusion.

**Appendix Figure A17:**
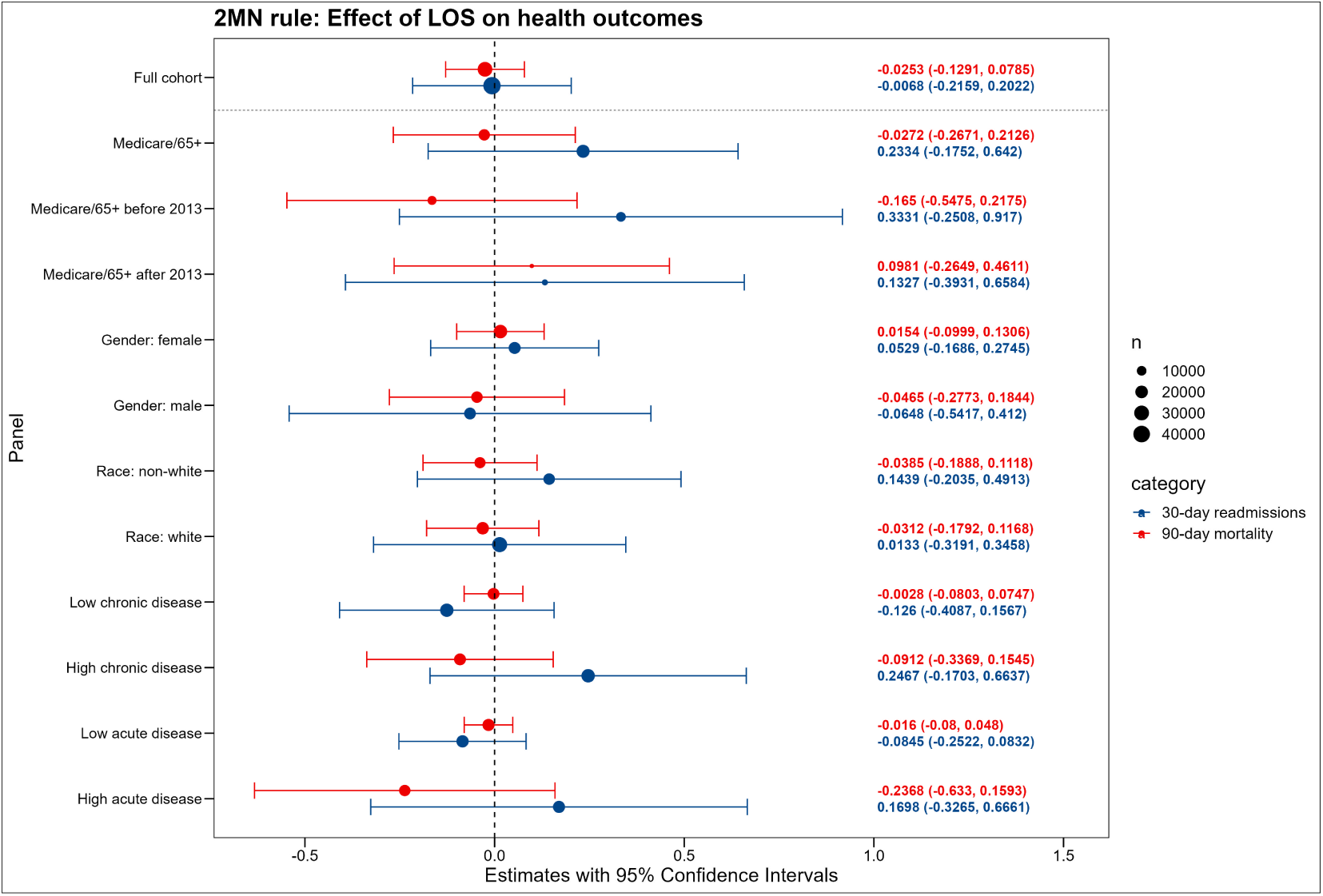
Effect of LOS on health outcomes for the 2MN rule sample *Note:* Estimated using local polyomial regression. Sample without data bunching exclusion.

**Appendix Figure A18:**
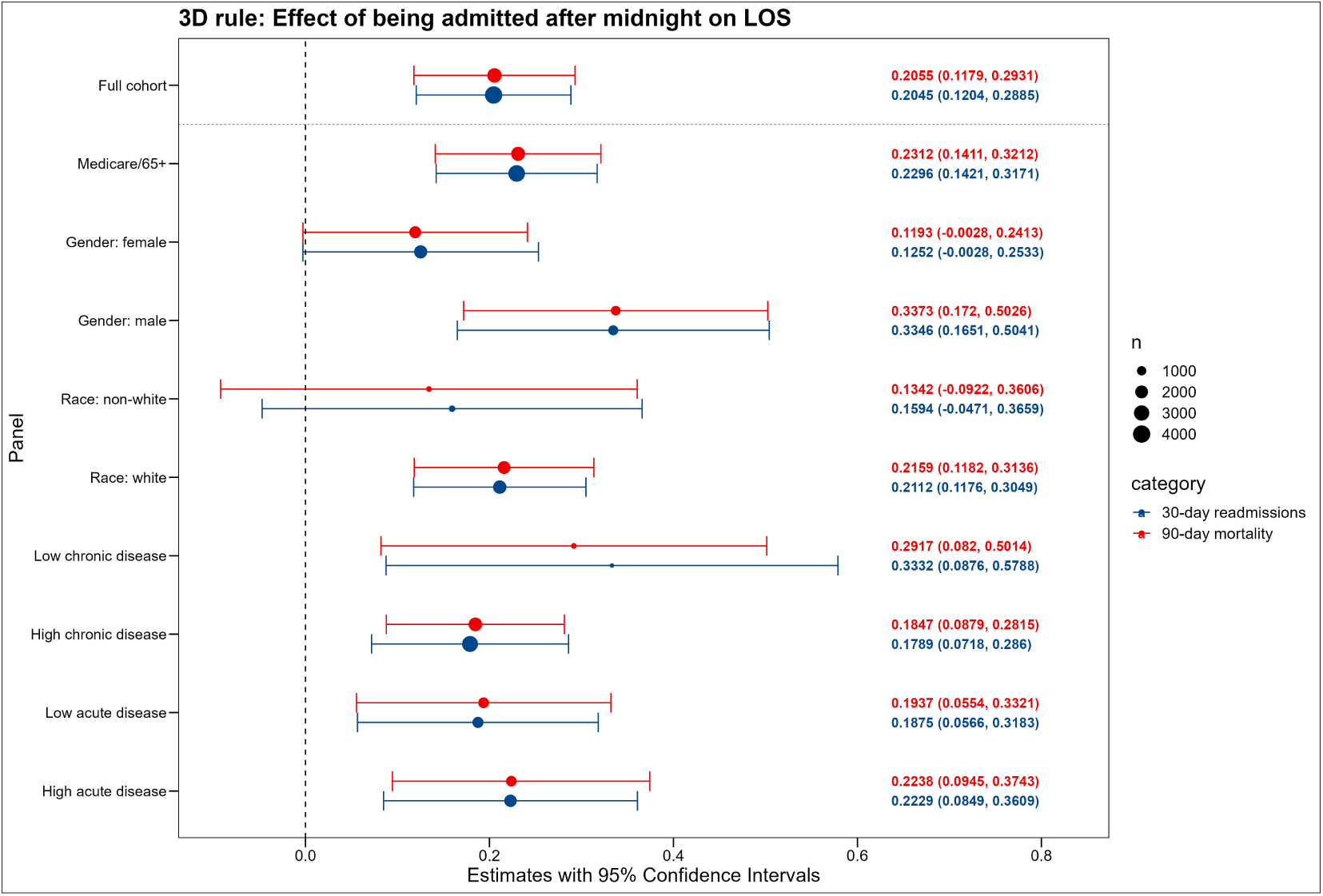
Effect of being admitted after midnight on LOS for the 3D rule sample *Note:* Estimated using local polyomial regression. Sample without data bunching exclusion.

**Appendix Figure A19:**
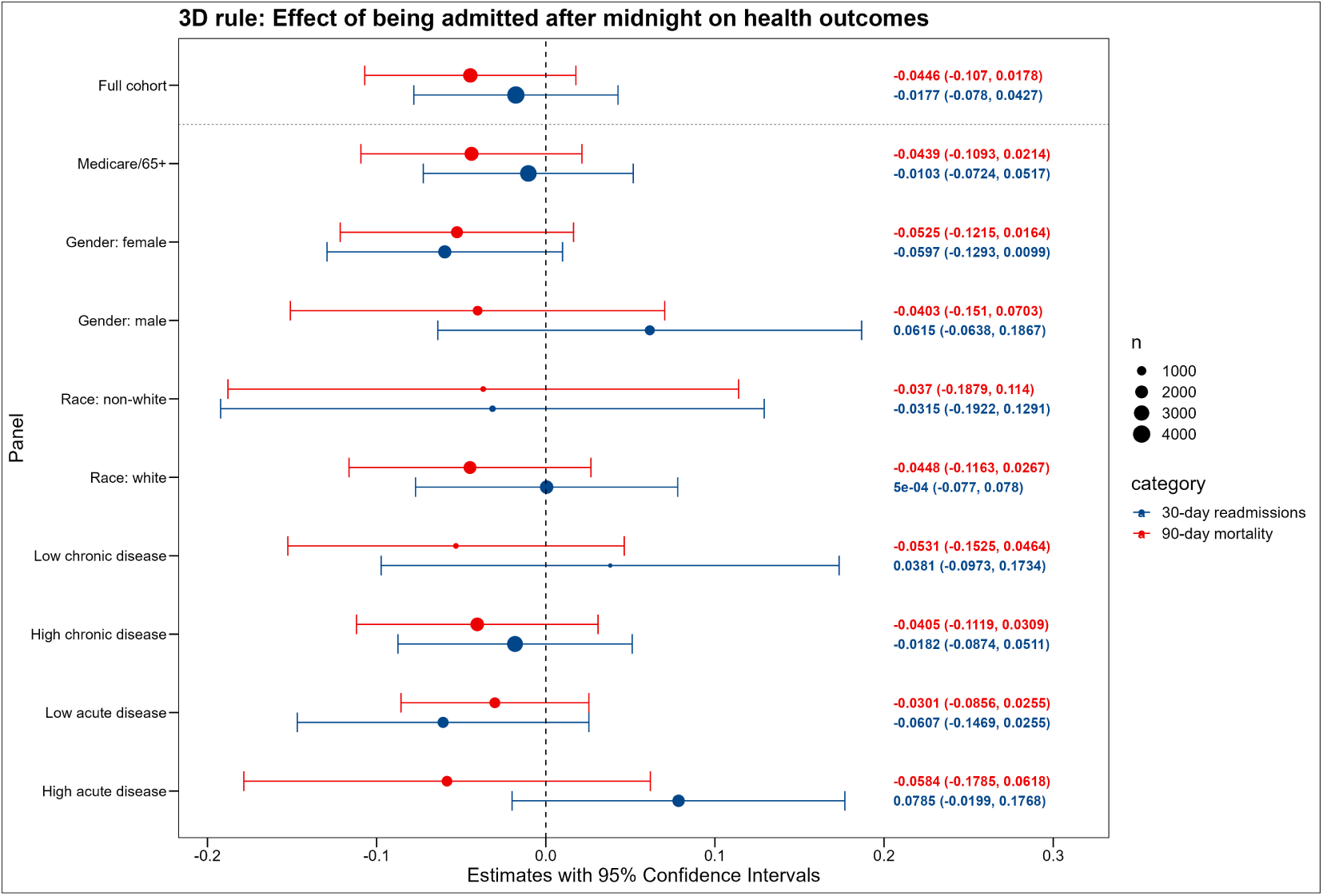
Effect of being admitted after midnight on health outcomes for the 3D rule sample *Note:* Estimated using local polyomial regression. Sample without data bunching exclusion.

**Appendix Figure A20:**
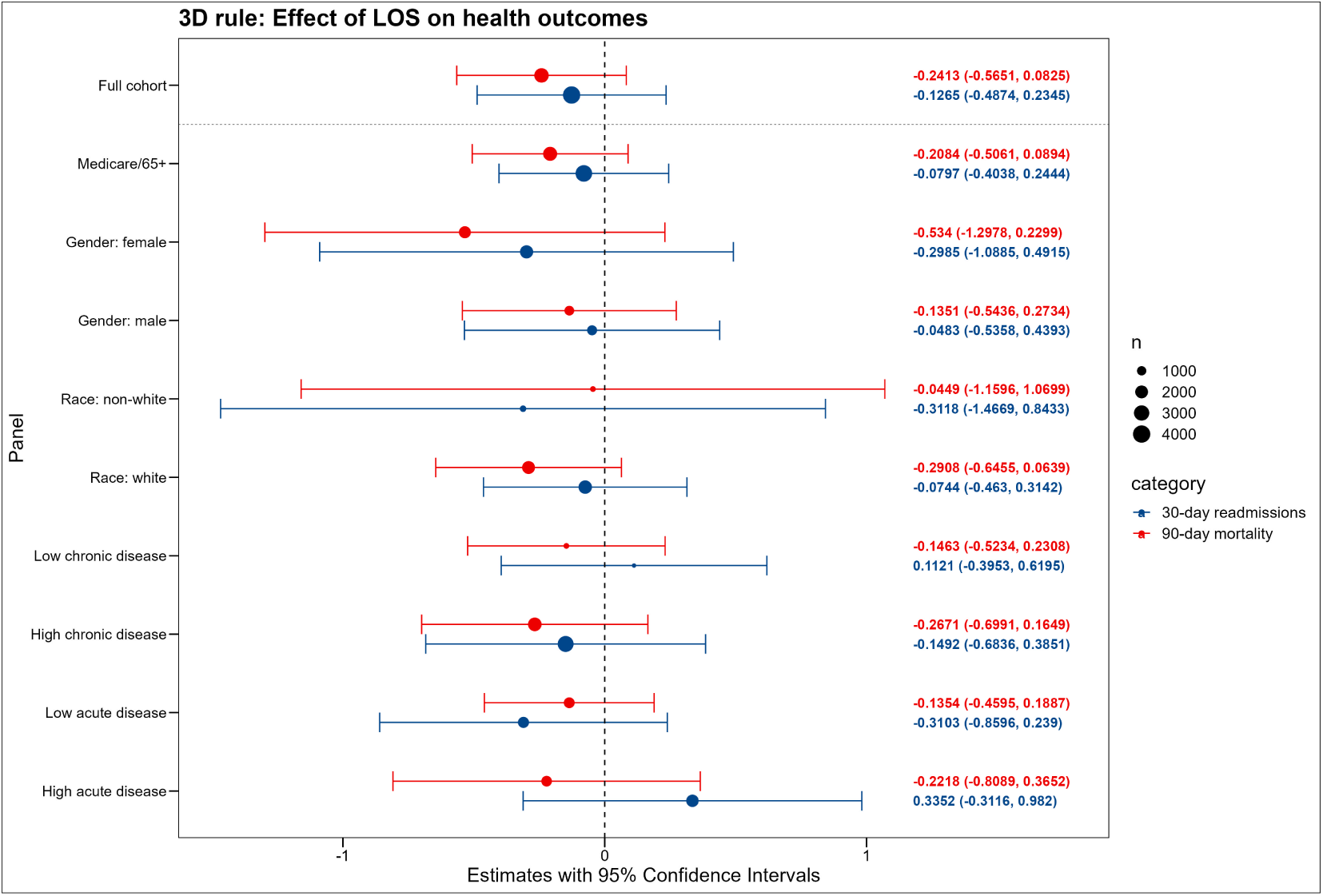
Effect of LOS on health outcomes for the 3D rule sample *Note:* Estimated using local polyomial regression. Sample without data bunching exclusion.

### B Tables

**Appendix Table A2:** Regression discontinuity estimates for covariates, 2MN rule.

| Variable | Estimate (regression discontinuity) | p-value |
| --- | --- | --- |
| Age | -0.02 | 0.44 |
| Gender | -0.01 | 0.55 |
| Race: white | 0.01 | 0.42 |
| Main language=English | -0.00 | 0.57 |
| Marital status: divorced | -0.02 | 0.06 |
| Marital status: married | 0.03 | 0.02 |
| Marital status: unknown | -0.00 | 0.41 |
| Marital status: single | -0.01 | 0.62 |
| Marital status: widowed | -0.00 | 0.81 |
| Insurance: Medicaid | -0.02 | 0.04 |
| Insurance: Medicare | -0.02 | 0.05 |
| Insurance: Other | 0.04 | 0.00 |
| Year group: 11-13 | 0.01 | 0.21 |
| Year group: 14-16 | 0.02 | 0.14 |
| Year group: 17-19 | -0.02 | 0.09 |
| Charlson wscore | -0.04 | 0.41 |
| Died in hospital | -0.00 | 0.74 |
| ICU stay | 0.01 | 0.44 |

**Appendix Table A4:** Compliers, always-takers and never-takers characteris- tics.

| Variable | All patients | Compliers | Always-takers | Never-takers |
| --- | --- | --- | --- | --- |
| Age | 58.90 | 62.85 | 63.42 | 53.65 |
| Gender: male | 0.49 | 0.49 | 0.48 | 0.50 |
| Charlson wscore | 1.85 | 2.51 | 2.67 | 1.02 |
| Acute mortality risk | 0.07 | 0.11 | 0.12 | 0.03 |
| **Insurance** |  |  |  |  |
| Medicaid | 0.11 | 0.08 | 0.08 | 0.13 |
| Medicare | 0.39 | 0.45 | 0.46 | 0.31 |
| Other | 0.51 | 0.47 | 0.46 | 0.55 |
| Language: English | 0.90 | 0.89 | 0.89 | 0.91 |

Appendix Table A4 – continued from previous page
| Variable | All patients | Compliers | Always-takers | Never-takers |
| --- | --- | --- | --- | --- |
| **Marital status** |  |  |  |  |
| Divorced | 0.08 | 0.08 | 0.08 | 0.07 |
| Married | 0.37 | 0.40 | 0.40 | 0.33 |
| Marital status unknown | 0.02 | 0.03 | 0.02 | 0.02 |
| Single | 0.42 | 0.36 | 0.35 | 0.49 |
| Widowed | 0.11 | 0.13 | 0.14 | 0.08 |
| **Race** |  |  |  |  |
| Native American | 0.00 | 0.00 | 0.00 | 0.00 |
| Asian | 0.03 | 0.03 | 0.03 | 0.03 |
| Black | 0.18 | 0.15 | 0.16 | 0.20 |
| Race unknown | 0.02 | 0.03 | 0.02 | 0.02 |
| White | 0.66 | 0.69 | 0.69 | 0.62 |
| **Year of admission** |  |  |  |  |
| Year 11-13 | 0.23 | 0.21 | 0.23 | 0.23 |
| Year 14-16 | 0.18 | 0.20 | 0.19 | 0.17 |
| Year 17-19 | 0.12 | 0.15 | 0.13 | 0.11 |
| **Type of admission** |  |  |  |  |
| Direct observation | 0.02 | 0.01 | 0.00 | 0.04 |
| Elective | 0.00 | 0.00 | 0.00 | 0.00 |
| Emergency | 0.48 | 0.68 | 0.68 | 0.28 |
| Urgent | 0.03 | 0.06 | 0.05 | 0.01 |
| **Admission location** |  |  |  |  |
| Ambulatory surgery | 0.00 | 0.00 | 0.00 | 0.00 |
| Clinical referral | 0.01 | 0.01 | 0.01 | 0.01 |
| Emergency room | 0.77 | 0.72 | 0.73 | 0.82 |
| Hospital transfer | 0.06 | 0.11 | 0.08 | 0.04 |
| Internal psych | 0.00 | 0.01 | 0.01 | 0.00 |
| Unknown | 0.00 | 0.00 | 0.00 | 0.00 |
| Pacu | 0.00 | 0.00 | 0.00 | 0.00 |
| Physician referral | 0.09 | 0.09 | 0.09 | 0.07 |
| Procedure site | 0.00 | 0.00 | 0.00 | 0.00 |
| SNF | 0.01 | 0.02 | 0.02 | 0.01 |
| Walk-in | 0.05 | 0.05 | 0.05 | 0.05 |
| **Discharge location** |  |  |  |  |
| Unknown | 0.35 | 0.05 | 0.05 | 0.64 |
| Acute hospital | 0.00 | 0.00 | 0.01 | 0.00 |
| Against advice | 0.01 |  | 0.01 | 0.01 |

Appendix Table A4 – continued from previous page
| Variable | All patients | Compliers | Always-takers | Never-takers |
| --- | --- | --- | --- | --- |
| Assisted living | 0.00 | 0.00 | 0.00 | 0.00 |
| Died | 0.02 | 0.03 | 0.02 | 0.01 |
| Home | 0.30 | 0.35 | 0.36 | 0.26 |
| Home care | 0.15 | 0.25 | 0.25 | 0.05 |
| Hospice | 0.01 | 0.01 | 0.02 | 0.00 |
| Long term care | 0.02 | 0.03 | 0.03 | 0.00 |
| Other | 0.00 | 0.01 | 0.01 | 0.00 |
| Psych | 0.01 | 0.01 | 0.01 | 0.00 |
| Rehab | 0.02 | 0.05 | 0.05 | 0.00 |
| SNF | 0.11 | 0.20 | 0.19 | 0.01 |

**Notes:** 95 percent confidence intervals in parentheses. Complier characteristics were derived based on the sample of patients without LOS restriction. Median LOS was 2.54 days.

**Appendix Table A5:**
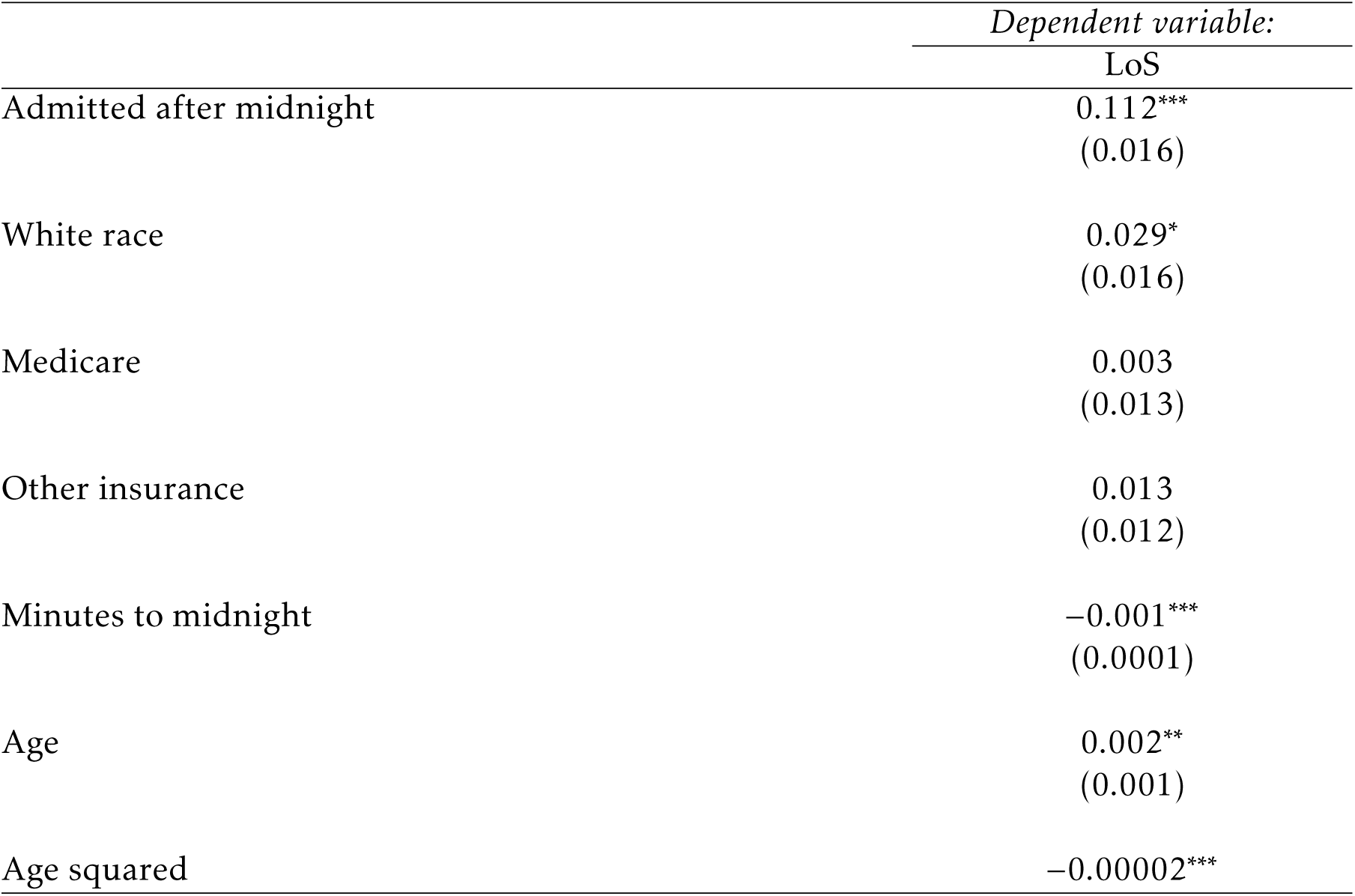
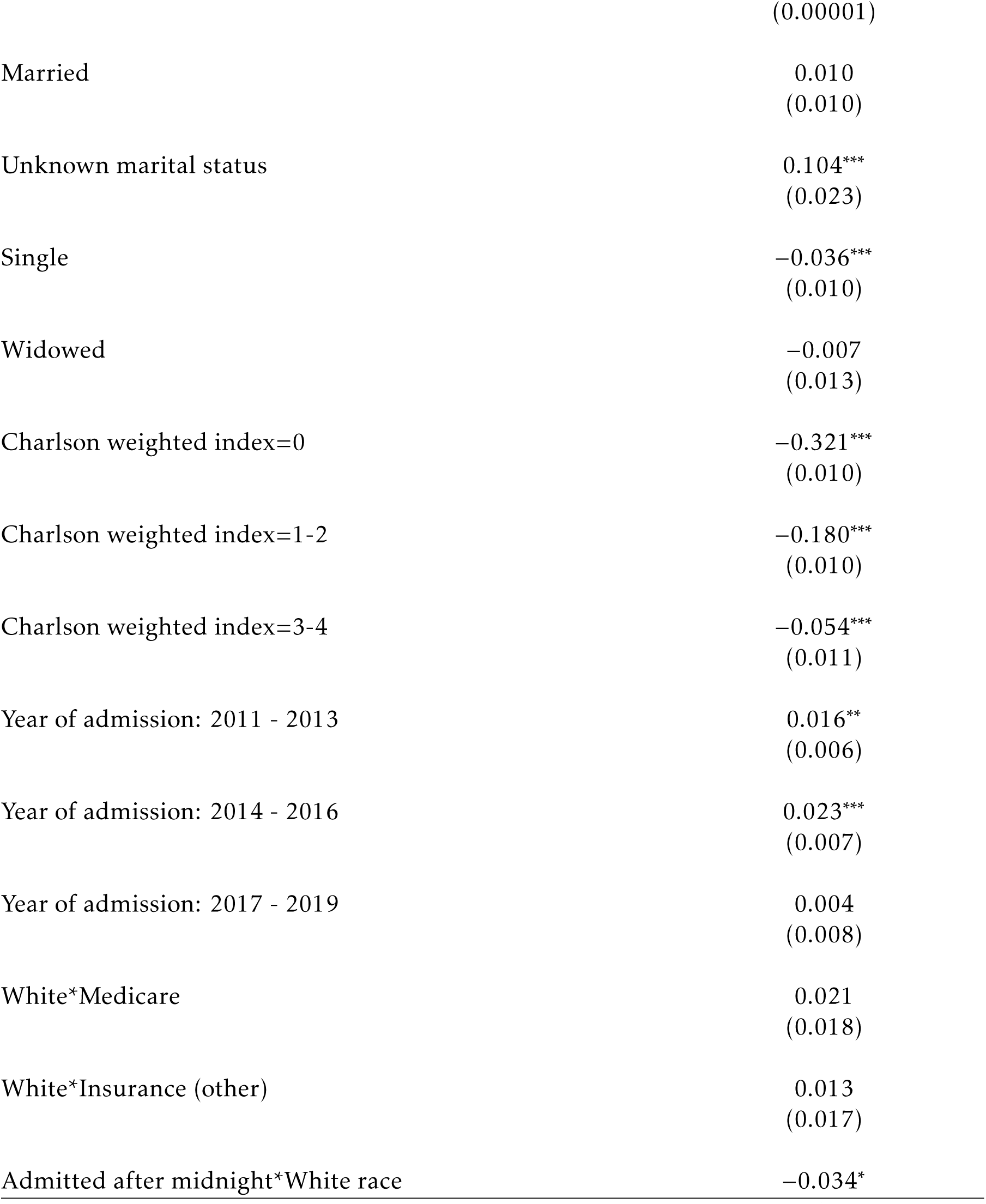
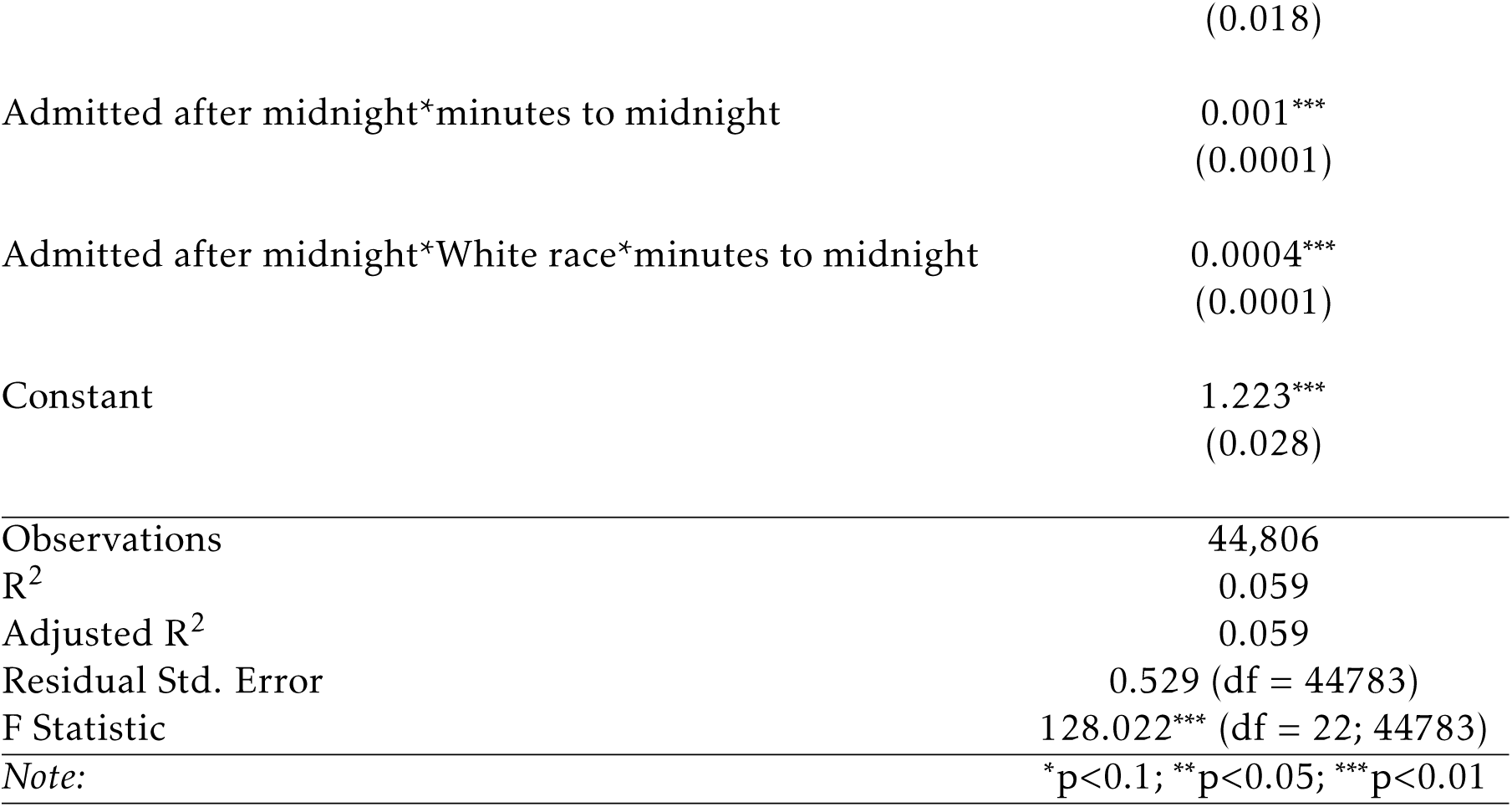
Regression Results for race (2MN rule)

**Appendix Table A6:** Regression results for gender (2MN rule)

|  | *Dependent variable:* |
| --- | --- |
|  | LoS |
| Admitted after midnight | 0.079***<br>(0.014) |
| White race | 0.054***<br>(0.016) |
| Medicare | 0.034***<br>(0.013) |
| Other insurance | 0.049***<br>(0.012) |
| Minutes to midnight | -0.001***<br>(0.0001) |
| Continued on next page |  |

**Table A6 continued from previous page**
|  | *Dependent variable:* |
| --- | --- |
|  | LoS |
| White race | 0.046***<br>(0.005) |
| Age | 0.002**<br>(0.001) |
| Age squared | −0.00002***<br>(0.00001) |
| Married | 0.012<br>(0.010) |
| Unknown marital status | 0.104***<br>(0.023) |
| Single | −0.035***<br>(0.010) |
| Widowed | −0.008<br>(0.013) |
| Charlson weighted index=0 | −0.322***<br>(0.010) |
| Charlson weighted index=1-2 | −0.181***<br>(0.010) |
| Charlson weighted index=3-4 | −0.054***<br>(0.011) |
| Year of admission: 2011 - 2013 | 0.016**<br>(0.006) |
| Year of admission: 2014 - 2016 | 0.023***<br>(0.007) |
| Year of admission: 2017 - 2019 | 0.004<br>(0.008) |

**Table A6 continued from previous page**
|  | *Dependent variable:* |
| --- | --- |
|  | LoS |
| White race*Medicare | -0.040**<br>(0.018) |
| White race*Other insurance | -0.060***<br>(0.017) |
| Admitted after midnight*White race | 0.021<br>(0.017) |
| Admitted after midnight*Minutes to midnight | 0.002***<br>(0.0001) |
| Admitted after midnight*White race*Minutes to midnight | -0.0003*<br>(0.0001) |
| Constant | 1.186***<br>(0.029) |
| Observations | 44,806 |
| R<sup>2</sup> | 0.059 |
| Adjusted R<sup>2</sup> | 0.059 |
| Residual Std. Error | 0.529 (df = 44782) |
| F Statistic | 122.870*** (df = 23; 44782) |
| *Note:* | *p<0.1; **p<0.05; ***p<0.01 |

**Appendix Table A7:** Regression Results for race (3D rule)

|  | *Dependent variable:* |
| --- | --- |
|  | LoS |
| Admitted after midnight | 0.183**<br>(0.073) |
| White race | 0.026<br>(0.117) |
| Medicare | 0.036<br>(0.089) |

**Table A7 continued from previous page**
|  | *Dependent variable:* |
| --- | --- |
|  | LoS |
| Other insurance | 0.075<br>(0.093) |
| Minutes to midnight | −0.001***<br>(0.0002) |
| Age | 0.004<br>(0.007) |
| Age squared | −0.00003<br>(0.00005) |
| Married | 0.036<br>(0.039) |
| Unknown marital status | −0.026<br>(0.067) |
| Single | 0.004<br>(0.039) |
| Widowed | 0.036<br>(0.040) |
| Charlson weighted index=0 | −0.040<br>(0.032) |
| Charlson weighted index=1-2 | −0.041<br>(0.027) |
| Charlson weighted index=3-4 | −0.022<br>(0.029) |
| Year of admission: 2011 - 2013 | 0.058**<br>(0.024) |
| Year of admission: 2014 - 2016 | 0.048*<br>(0.028) |

**Table A7 continued from previous page**
|  | *Dependent variable:* |
| --- | --- |
|  | LoS |
| Year of admission: 2017 - 2019 | 0.048<br>(0.033) |
| White race*Medicare | 0.011<br>(0.119) |
| White race*Other insurance | 0.016<br>(0.124) |
| Admitted after midnight*White race | 0.009<br>(0.078) |
| Admitted after midnight*Minutes to midnight | 0.001<br>(0.001) |
| Admitted after midnight*White race*Minutes to midnight | 0.001<br>(0.001) |
| Constant | 2.973***<br>(0.272) |
| Observations | 2,913 |
| R<sup>2</sup> | 0.034 |
| Adjusted R<sup>2</sup> | 0.027 |
| Residual Std. Error | 0.510 (df = 2890) |
| F Statistic | 4.636*** (df = 22; 2890) |
| *Note:* | *p<0.1; **p<0.05; ***p<0.01 |

**Appendix Table A8:** Regression Results for gender (3D rule)

|  | *Dependent variable:* |
| --- | --- |
|  | LoS |
| Admitted after midnight | 0.191***<br>(0.057) |
| White race | -0.053 |

**Table A8 continued from previous page**
|  | *Dependent variable:* |
| --- | --- |
|  | LoS |
|  | (0.116) |
| Medicare | 0.029<br>(0.085) |
| Other insurance | 0.044<br>(0.088) |
| Minutes to midnight | −0.001***<br>(0.0002) |
| White race | 0.062***<br>(0.024) |
| Age | 0.004<br>(0.007) |
| Age squared | −0.00003<br>(0.00005) |
| Married | 0.033<br>(0.039) |
| Unknown marital status | −0.027<br>(0.067) |
| Single | 0.003<br>(0.039) |
| Widowed | 0.039<br>(0.040) |
| Charlson weighted index=0 | −0.038<br>(0.033) |
| Charlson weighted index=1-2 | −0.038<br>(0.027) |
| Charlson weighted index=3-4 | −0.021 |

**Table A8 continued from previous page**
|  | *Dependent variable:* |
| --- | --- |
|  | LoS |
|  | (0.029) |
| Year of admission: 2011 - 2013 | 0.057**<br>(0.024) |
| Year of admission: 2014 - 2016 | 0.047*<br>(0.028) |
| Year of admission: 2017 - 2019 | 0.046<br>(0.033) |
| White race*Medicare | 0.029<br>(0.117) |
| White race*Other insurance | 0.078<br>(0.122) |
| Admitted after midnight*White race | -0.003<br>(0.067) |
| Admitted after midnight*Minutes to midnight | 0.001**<br>(0.001) |
| Admitted after midnight*White race*Minutes to midnight | 0.0001<br>(0.001) |
| Constant | 2.964***<br>(0.268) |
| Observations | 2,913 |
| R<sup>2</sup> | 0.034 |
| Adjusted R<sup>2</sup> | 0.026 |
| Residual Std. Error | 0.510 (df = 2889) |
| F Statistic | 4.390*** (df = 23; 2889) |
| *Note:* | *p<0.1; **p<0.05; ***p<0.01 |

**Appendix Table A3:** Regression discontinuity estimates for covariates, 3D rule.

| Variable | Estimate (regression discontinuity) | p-value |
| --- | --- | --- |
| Age | 0.04 | 0.61 |
| Gender | 0.01 | 0.86 |
| Race: white | 0.03 | 0.49 |
| Main language=English | 0.03 | 0.50 |
| Marital status: divorced | -0.01 | 0.79 |
| Marital status: married | -0.05 | 0.37 |
| Marital status: unknown | -0.00 | 0.90 |
| Marital status: single | -0.03 | 0.60 |
| Marital status: widowed | 0.08 | 0.04 |
| Insurance: Medicaid | -0.02 | 0.34 |
| Insurance: Medicare | 0.05 | 0.31 |
| Insurance: Other | -0.05 | 0.36 |
| Year group: 11-13 | -0.05 | 0.22 |
| Year group: 14-16 | 0.02 | 0.62 |
| Year group: 17-19 | -0.00 | 0.91 |
| Charlson wscore | -0.19 | 0.45 |
| Died in hospital | 0.00 | 1.00 |
| ICU stay | 0.04 | 0.30 |

### C Additional text

In fuzzy RD, an individual that receives the treatment (i.e., an extra day in hospital) as a consequence of being on the “treatment side” of the cut-off (i.e., admitted after midnight), and otherwise would not have taken the treatment (i.e., would have had a day less in hospital if being admitted before midnight) is referred to as complier [31]. Other than that, patients can be either “never-takers” and “always-takers”.

While never-takers are patients that never stay an extra day in hospital, even when admitted after midnight, always-takers always stay an extra day in hospital, irrespective of the time of admission. Therefore, they are not affected by the instrument [31]. While it cannot be observed whether an individual falls into the category of complier, never-taker or always-taker, the group characteristics of compliers can be obtained by deriving

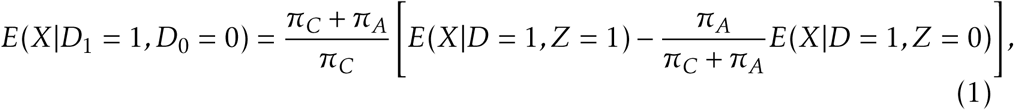

where *π_C_* indicates the fraction of compliers, *π_A_* the fraction of always-takers, *D* is a dummy that is equal to 1 if the individual had a long hospital stay and 0 if the individual had a short hospital stay, and *Z* is a dummy equal to 1 if an individual was admitted after midnight and 0 if admitted before midnight [15]. Thus, the complier characteristics are easy to obtain using observable sample means. However, since our main treatment variable, the LOS, is continuous, we cannot derive the complier characteristics based on this variable without further transformation. Since a binary variable is required (see *D* in formula 1), we transform our otherwise continuous LOS into a binary variable that is equal to 1 if a patient had a LOS of above median, and 0 otherwise.

#### Bandwidth Choice

While setting the bandwidth too broad, the validity of the RD design may decrease as the groups may differ in characteristics. If we set the bandwidth too narrow, statistical power decreases. To achieve an optimal bandwidth, the approach of Calonico et al. 44 as built into the rdrobust package [32] is straightforward. This approach allows for robust bias-corrected confidence intervals with minimal coverage error, which makes this approach superior to mean-squared error optimized bandwidth (see e.g. Imbens and Kalyanaraman 45).

#### Results: Complier characteristics

As explained earlier, fuzzy RD estimates hold only for the group of compliers, a group of patients which typically differ from the general population. Hence, we estimated complier characteristics, as described in the methods section, in order to identify how compliers differ from the general study population, never-takers and always-takers.

Compared to the sample average, compliers had higher average age and higher chronic comorbidity as indicated by the Charlson weighted score, compared to the sample average. They were further more likely to be covered by Medicare as opposed to Medicaid or other insurance, and were less likely to be single but slightly more likely to be married. Differences in race were small, with compliers being slightly more likely to be white and less likely to be black. Further, they were more likely to be admitted as emergency or urgent case, and more likely to be admitted from another hospital. Also, they were less likely to be discharged to an unknown location, and more likely to be discharged to home care or home (Table A4).

#### Robustness check

We performed several additional analyses to test the robustness of our results. First, in an analysis that included patients admitted in the +/-6 minutes surrounding midnight, findings were similar in both the 2MN rule and 3D rule cohorts, suggesting against manipulation of documentation for admission times occurring around midnight that would bias results (Fig. A15, A16, A17, A18, A19, A20). Second, when we restricted our sample to patients with LOS *>* 4.5 days, there was no discontinuous difference in outcomes surrounding the midnight boundary (for details see Appendix C), suggesting against effects of being admitted around midnight on LOS that aren’t related to the Medicare rules.

#### Patients beyond 4.5 days of LOS

We further checked whether there remains a discontinuity for patients with LOS *>*4.5 days, since this would indicate that we missed a policy leading to a discontinuity for patients with long stays, or that something with our approach would be fundamentally wrong. Contrary, if we do not observe a midnight-discontinuity for patients with LOS *>*4.5 days, we may consider that as strong evidence for our reasoning for the two policies under consideration to be the true drivers of the discontinuity. Graphical analysis and statistical analysis indicated that there is no discontinuity for patients with LOS *>*4.5 days (Fig. A14).

#### Gender and race subgroup results

While it seems especially reasonable to not delay discharge for patients with high chronic/acute disease, it was puzzling to find differences based on gender and race, where especially the differences regarding race were stark. This result raised the need for further investigation, especially since discrimination based on gender or race are known from other contexts in healthcare [33, 34, 35, 36].

One plausible explanation may be that females and non-white patients were different regarding their insurance type, thus yielding different incentives for being affected by the respective rules. For the 2MN rule, we found that white patients were more likely to be insured by Medicare as opposed to non-white patients, potentially explaining differences (*White*: Medicaid 6.67%; Medicare 40.1%; other: 53.2%; *non-White*: Medicaid 16.5%; Medicare: 28.1%; other: 55.4%).

To get a better understanding of the finding, we run additional analysis using interaction terms to identify whether differences may be explained by covariates such as type of insurance coverage. The regression analysis revealed a significant interaction between race and being admitted after midnight at the 10% significance level, whereby white patients had a shorter stay than non-white patients, even after controlling for insurance type and respective interactions (Table A5). This result warrants further investigation as to why non-white patients tend to be more affected by the 2MN rule even after controlling for insurance effects (despite the limitation that we do not have sufficiently precise data on Medicare advantage).

For gender, we found a relatively similar insurance breakdown for males and females (*males*: Medicaid 10.5%; Medicare 34.5%; other 54.9%; *females*: Medicaid 9.76%; Medicare 37.2%; other 53.1%). Running the interaction-term regression, we found that the interaction between gender and being admitted after midnight was insignificant. Therefore, the subgroup difference for gender seems to be mediated by control variables, not pointing towards discrimination regarding the 2MN rule effect. Independently, female gender itself was associated with increased LOS (Table A6).

For the 3D rule, we also found that patients with low chronic disease experienced a starker effect of the discontinuity on LOS. Further, we found the opposite with respect to gender and race: female patients and white patients experienced longer LOS as compared to their counterparts (Fig. A8). As for the 2MN rule, insurance type distribution may be a plausible explanation. As we analyzed the coverage within the white vs. non-white population, we indeed found that while insurance coverage by Medicare accounted for 78.8% of hospital stays, for non-white patients, only 64.5% of patients were covered by Medicare. For non-whites, the percentage of Medicaid and “other” (including Medicare advantage) was larger than for whites (Medicaid: 6.89% vs. 1.97%; “other”: 28.6% vs. 19.30%) for the 3D rule affected population, potentially explaining (part of) the disparities.

Interaction-term regression analysis for the 3D rule identified no significant association between the interaction of race and insurance type and LOS, nor was race itself associated with LOS. Hence, the difference in effect size for the subsample by race may have been explained by factors we controlled for such as insurance type. Further, for gender, we found no significant effect of the interaction of gender and being admitted after midnight on LOS, nor an effect of the variable gender itself after controlling for covariates (Table A6; Table A7).

## Footnotes

1 For OLS, the effect of LOS on 30-day readmissions appeared statistically significant in low acute disease patients, suggesting that an extra day spent in the hospital may be beneficial for these patients. However, since OLS is more susceptible to functional form bias, we assume that the local polynomial estimates are more accurate

